# Network-based drug repurposing for psychiatric disorders using single-cell genomics

**DOI:** 10.1101/2024.12.01.24318008

**Authors:** Chirag Gupta, Noah Cohen Kalafut, Declan Clarke, Jerome J. Choi, Kalpana Hanthanan Arachchilage, Saniya Khullar, Yan Xia, Xiao Zhou, Mark Gerstein, Daifeng Wang

## Abstract

Neuropsychiatric disorders lack effective treatments due to a limited understanding of underlying cellular and molecular mechanisms. To address this, we integrated population-scale single-cell genomics data and analyzed cell-type-level gene regulatory networks across schizophrenia, bipolar disorder, and autism (23 cell classes/subclasses). Our analysis revealed potential druggable transcription factors co-regulating known risk genes that converge into cell-type-specific co-regulated modules. We applied graph neural networks on those modules to prioritize novel risk genes and leveraged them in a network-based drug repurposing framework to identify 220 drug molecules with the potential for targeting specific cell types. We found evidence for 37 of these drugs in reversing disorder-associated transcriptional phenotypes. Additionally, we discovered 335 drug-associated cell-type eQTLs, revealing genetic variation’s influence on drug target expression at the cell-type level. Our results provide a single-cell network medicine resource that provides mechanistic insights for advancing treatment options for neuropsychiatric disorders.

## Introduction

Psychiatric disorders, including schizophrenia (SCZ), bipolar disorder (BPD), and autism spectrum disorder (ASD) are complex conditions lacking effective treatments due to complex gene regulation and cellular heterogeneity(*1*). Single-cell multi-omics offer a promising avenue for analyzing dysregulated epigenomic and transcriptional states and identifying specific cell types affected by the disease(*2–5*). Analysis of cell type gene regulatory networks and dysregulated pathways that underpin disease can lead to identifying novel therapeutic targets for specific cell types(*6*, *7*). This resolution enables drug repurposing efforts to focus on modulating abnormalities in gene regulation within specific cell types, thereby facilitating the discovery of effective treatments that are tailored to the underlying etiology of each disease. However, post-genomic drug repurposing efforts for cell types perturbed in neuropsychiatric disorders remain poorly characterized, mainly due to the lack of uniformly processed datasets that allow investigation across disorders using control and disease groups.

This gap was recently filled by the PsychENCODE consortium(*8*). The Phase II efforts of the consortium generated single-cell sequencing data from adult brains with multiple neuropsychiatric disorders from the human prefrontal cortex. The collection provides uniformly processed single-nucleus (sn) RNA-seq and snATAC-seq, and snMultiome. The dataset comprises >2.8 M nuclei across 28 distinct brain cell types from 388 individual brains. The dataset accompanies 24 cell type gene regulatory networks (GRN) inferred using data from healthy donors. The cell type GRNs were found to be significantly different between cell types, with extensive rewiring and differential usage of enhancers and promoters by the same TFs. The study also highlights interesting patterns, such as co-regulation of disease risk genes in a cell type specific manner. These previous observations invite a more thorough, cross-disorder GRN comparison between cell types to highlight key differences in the regulatory dynamics of psychiatric disorders, as these lines of investigation had not been explored in the original PEC2 work. The PEC2 dataset also presents a unique opportunity to test the feasibility of repurposing drugs to target specific cell clusters implicated in neuro-psychiatric diseases.

GRNs offer a powerful means of linking promoters and enhancers with interactions between genes and their regulators (such as TFs). TFs effectively act as molecular “switches” that control the transcriptional output of cells and play important roles in disease phenotypes(*9*), thereby making them promising therapeutic targets(*9*). Recent advances in ligand engineering have challenged the notion of TFs as being ‘undruggable’, a perspective that can mainly be attributed to their structural and biophysical complexity(*10–12*). TFs can be interesting targets for drugs designed to restore transcriptional phenotypes(*13*), selectively repress or activate disease regulons (gene sets under similar regulatory control)(*14–16*), or when combining two or more drugs with different mechanisms to boost clinical efficacy(*17*, *18*). While there are several approaches for using genomic data for drug repurposing(*19*), network medicine-based strategies have been widely adopted for brain-related(*20–23*) and other human(*24–26*). The strategy aims to test the network proximity between disease gene sets and drug targets in biological networks such as via protein-protein interactions, thus increasing the likelihood that they affect the disease through multiple network pathways. Thus, the availability of population-scale single-cell sequencing data can enhance network medicine-based strategies by revealing key dysregulated mechanisms at the cell type level. Furthermore, an integrated analysis of single-cell multi-omics, genetic variants, and drug targets can be beneficial for applications in personalized medicine.

However, the identification of effective drug targets for brain disorders remains hindered by limited knowledge of disease-risk genes and their cell-type specificity. Many disease genes may be coregulated by similar regulatory mechanisms(*27–29*), suggesting that cell-type GRNs contain the information necessary to bridge the gap between known risk genes and novel candidate genes. Graph learning approaches, which leverage network information to identify similar nodes and prioritize edges, have successfully inferred disease genes in various networks e.g., protein-protein interactions(*30–33*). However, the application of graph learning to cell-type GRNs for inferring novel candidate disorder genes, especially at the cell type level, remains largely underexplored. Novel risk genes with cell type specificity can help identify relevant drug targets.

To the best of our knowledge, there are no previous estimates of repurposable drug candidates for neuropsychiatric disorders at the cell type level, despite their increasing importance in pharmaceutical development(*7*). To address this gap, our study explores a single-cell network medicine-based approach to drug repurposing for three neuropsychiatric disorders: ASD, SCZ, and BPD. Our study is divided into four main parts. First, we infer cross-disorder cell type GRNs by integrating snRNA-seq data and scATAC-seq single-cell data from a population of 140 diagnosed individuals and 107 age-matched controls in the PEC2 cohort. We then survey network properties that characterize these GRNs and identify differential TF regulons enriched with drug targets. Second, we utilize the GRNs to train graph neural networks to predict cell type specific disorder risk genes. Third, we leverage these prediction models to identify and prioritize drugs that could potentially restore the transcriptional phenotype of neuropsychiatric disorders to a healthy state. Finally, we identify genetic variants that explain intrinsic expression variation of repurposable drug targets in an attempt to find drug-associated cell type eQTLs. Our study allowed us to explore the potential genetic associations with approved drugs in complex brain diseases and traits, thereby contributing to the foundation of precision medicine.

### Cell-type gene regulatory network construction for drug repurposing in psychiatric disorders

We leverage the recently published PsychENCODE2 dataset for network-based drug repurposing to study psychiatric disorders (**Fig. S1**). This cross-cohort, uniformly processed dataset enables us to predict high-quality cell type GRNs across three psychiatric disorders: ASD, SCZ, and BPD **(Fig. 1A; see supplementary section 1)**. We utilize >1.64 million cells from 247 donors and apply a GRN inference pipeline to comprehensively analyze dysregulated transcriptional pathways, drug repurposing, and pharmacogenomics across 23 cell types and the three psychiatric disorders **(Fig. 1, S2A)**. We identify rewired regulatory genes that are influenced by specific drug treatments and patterns of co-regulated disorder risk genes and their convergence into modules **(Fig. 1B; see method details in supplementary section 2)**. We also utilize a machine learning framework that predicted 249 novel cell type disorder risk genes **(Fig. 1C; see method details supplementary section 3)**. Furthermore, using the cell type disorder GRNs and predicted disorder genes, we test a library of >800 drug molecules for their proximities to each disorder within each cell type. Our framework nominates 220 drugs, 38 of which have existing evidence for the potential to modulate the transcriptional phenotype of the disorders **(Fig. 1D; see method details supplementary section 4)**. This approach enables us to link prioritized drugs to individual donors based on their genotypes, allowing for the identification of drug-QTLs with applications in pharmacogenomics and precision medicine **(Fig. 1E; see method details supplementary section 5)**. We have made our analyses available on a web application hosted at https://daifengwanglab.shinyapps.io/psyDGN/.

**Figure 1:**
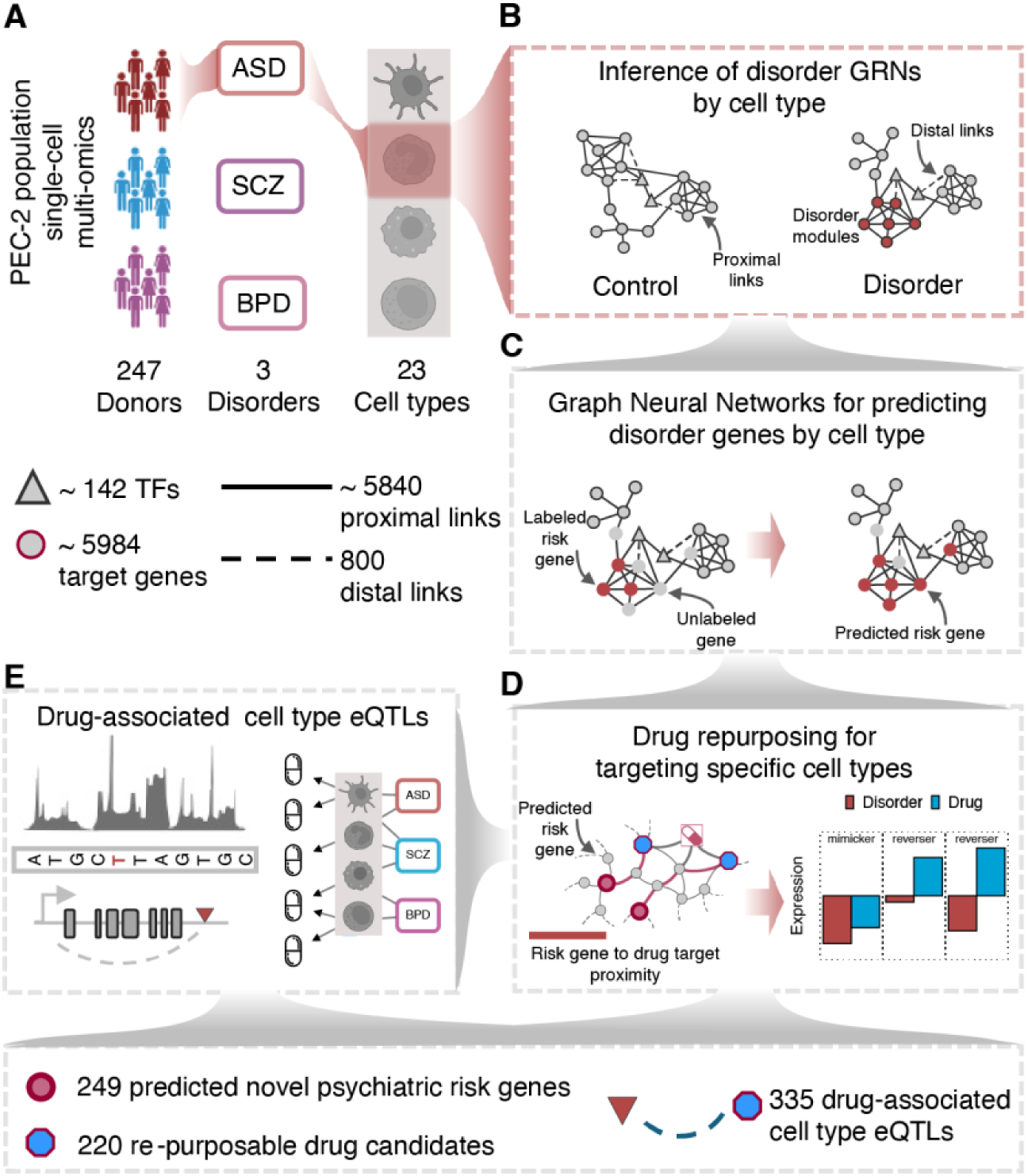
Network-based drug repurposing workflow. **A)** The PEC2 dataset was utilized to build gene regulatory networks for three psychiatric disorders across 23 cell types of the human brain. **B)** The GRNs link TFs to target genes via proximal interactions (from snRNA-seq) or distal interactions (from scATAC-seq) under both control (healthy) and disorder conditions from age-matched donor groups. The GRNs are then analyzed to identify druggable regulons and target gene modules that show strong statistical enrichment of known risk genes. **C)** We trained several variants of supervised Graph Neural Networks (GNNs) to predict and rank novel risk genes in a cell type specific manner. **D)** A network-proximity based procedure was applied to probe drugs with targets in close vicinity to risk genes predicted by the GNNs. **E)** Identification of QTLs that affect drug target gene expression.

### Druggable cell-type transcription factors for psychiatric disorders

We integrate scATAC-seq and snRNA-seq data to predict cell type GRNs in ASD, SCZ, and BPD, as well as in their respective control (healthy) individuals (**see Supplementary Note 1.2**). Briefly, we link transcription factors (TFs) to their potential target genes based on the strength of coexpression observed in the snRNA-seq data, thereby identifying proximal links. Additionally, we connect TFs to interacting enhancers and promoters in the scATAC-seq data to derive potential distal links (**Fig. 2A**). Overall, we predict GRNs across eight major cell types and 23 subclasses (see Methods). Overall, we infer 69 GRNs, which include an average 142 TFs and 5000 target genes linked via an average of 5840 proximal and 800 distal links (**Fig. S2B,C**). Cross-cohort analysis shows strong concordance between cell type GRNs (**Fig. S3**), suggesting the GRNs are insensitive to sampling bias.

**Figure 2:**
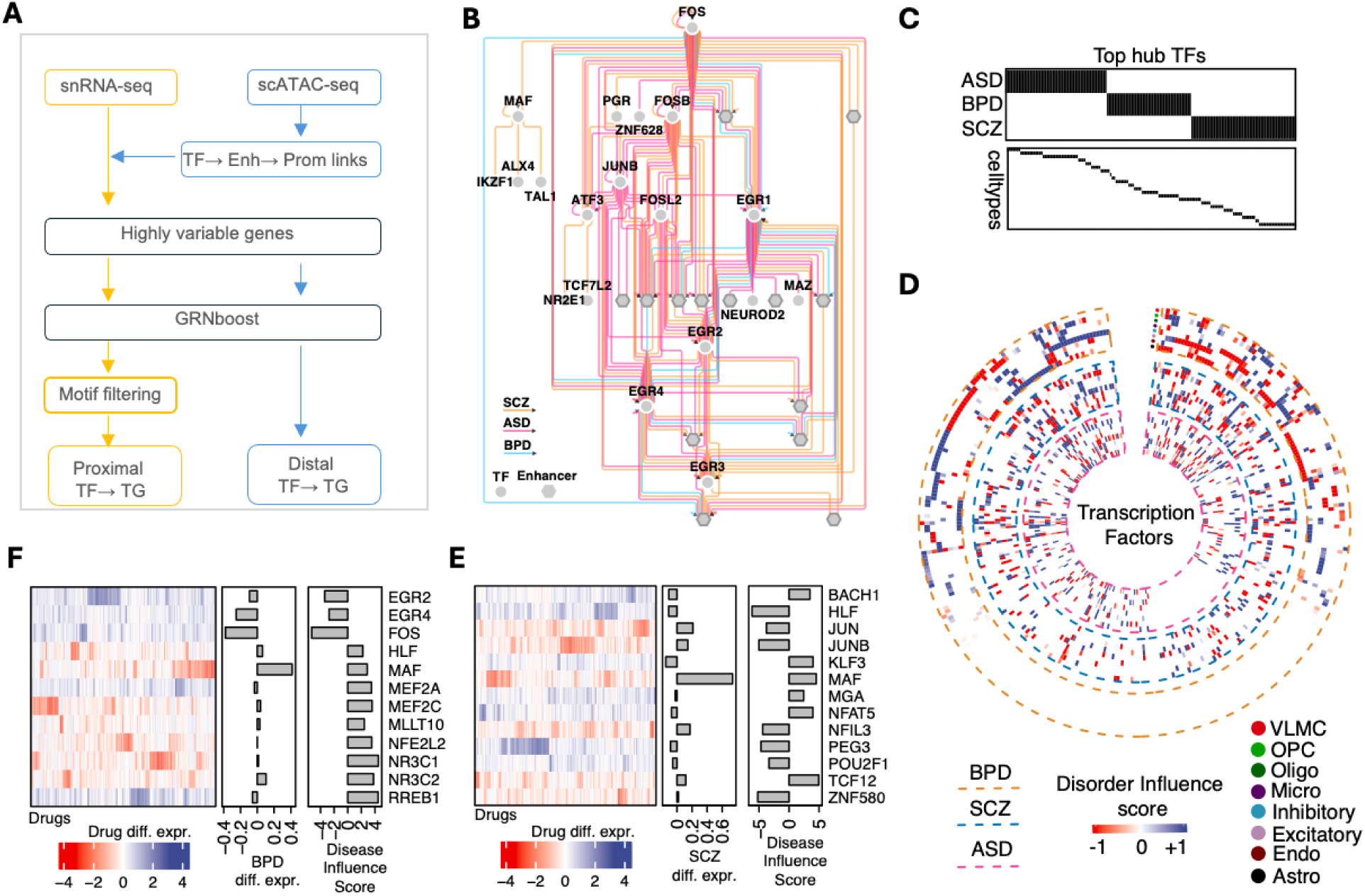
Druggable cell type transcription factors. **A)** Pipeline to integrate snRNA-seq and scATAC-seq data to predict TF→target gene links. **B)** An excitatory neuron sub-network plot showing differential usage of target genes by TFs in the different psychiatric disorders. **C)** TFs (x-axis) that act as hubs in cell types (x-axis, bottom panel) and disorder (x-axis top panel) GRNs. TF names are hidden for ease in visualization. **D)** Circular heatmaps showing the distribution of TF disease influence (DI) scores across cell types and disorders. The outermost, middle, and inner rings show distributions in bipolar disorder (BPD), schizophrenia (SCZ), and autism (ASD), respectively. The names of cell types along the x-axis and TFs along the y-axis are hidden for ease in visualization (the full version is shown in Fig. S5). The cell types of the heatmap are colored along a blue-to-red gradient representing gain or loss in influence, respectively. Only the outer ring of the heatmap is clustered; the inner two rings remain unclustered to preserve order. **E-F)** TFs with the largest changes in DI scores are shown on the y-axis along with their DI scores (right barplot) and log of change in expression in disorder with respect to control (left barplot). The heatmaps show a change in the expression of TFs in CNS cells treated with drug compounds (x-axis; names hidden) represented along a blue-to-red gradient indicating upregulation or downregulation, respectively.

We find extensive rewiring of TF regulons (the set of predicted target genes for a TF) between disorders (**Fig. 2B**). For instance, the FOS TF, a critical regulator of neural activity, exhibits differential usage of enhancers and promoters across different disorders (**Fig. 2B**). We generate a list of cell type and disorder-specific hubs (TFs with a disproportionately large number of targets) and bottlenecks (TFs that act as bridges) (**Fig. 2C, S4A,B; Supplementary Data S1**). The bottleneck TFs are those with a central role in connecting different parts of the network, measured by ’betweenness centrality.’ This score indicates which TFs are most influential in regulating other genes. To understand how these TFs change in different disorders, we compared their centrality scores in healthy (control) versus disorder networks, creating a ’Disorder Influence’ (DI) score. This DI score shows how much influence a TF gains or loses in each disorder (**see Supplementary Note 2.1**). The DI score reveals distinct cell type specific patterns in our GRNs (**Fig. 2D, S5**). We find excitatory neurons as the most perturbed cell type in ASD and BPD, whereas microglia are most perturbed in SCZ (**Fig. S4C**). Thus, the DI score helped us narrow down a list of the most rewired TFs for further interrogation (**Supplementary Data S2**).

Since we are interested in using GRNs for drug repurposing, we next investigate the distribution of drug targets along the distal links, as one proposed approach to include TFs into the druggable genome is to interrupt their binding sites on enhancers(*34*). Interestingly, we find a relatively over-enrichment of distally regulated drug targets in inhibitory neuronal cells of ASD individuals compared to the other two disorders (**Fig. S2D**). Furthermore, since TFs are not readily available as candidates for drug targets, we hypothesize that certain drug compounds may confer an indirect effect on TFs influencing its expression activity that can be monitored in disease cells. Indeed, we find several examples of TFs with high DI scores whose expression activity in cell lines treated with certain drug compounds is opposite to that of observed expression in the three disorders we studied **(Fig. 2E, 2F, Fig. S4D; Supplementary Data S3).**

Overall, our GRNs show extensive rewiring of TF nodes with differential usage of target genes. Our GRNs suggest a complex and heterogeneous landscape of gene regulation in psychiatric disorders, where different cell types and TFs may contribute uniquely to disease etiology. Understanding these distinct regulatory mechanisms at the level of target genes can provide a more holistic perspective into the molecular underpinnings of these disorders and potentially inform more targeted therapeutic strategies.

### Disorder risk genes converge on co-regulated gene modules

Since most known disease genes (and drug targets) are non-TF genes, it is imperative to analyze the GRNs at the target gene level to fully capture the regulatory dynamics and therapeutic potential of GRN models. We transform our directed cell type GRNs to undirected co-regulatory networks, which connect target gene pairs that have high overlaps between their regulators (**see Supplementary Note 3.1**). We then cluster these networks to derive modules of highly co-regulated genes, yielding >4000 dense modules across cell types and disorder networks (**Fig. S6A; Supplementary Data S4**), several of them enriched with gene ontology terms representing biological processes (**Fig. S6B,C; Supplementary Data S5**).

We then assessed whether these gene modules are preserved between control and disorder networks, using normalized mutual information (NMI) to compare clustering patterns. NMI values range from 0 (dissimilar) to 1 (similar), allowing us to identify distinct cell-type-specific patterns. For instance, modules in inhibitory neurons remain relatively unchanged in ASD and SCZ but exhibit drastic changes in BPD **(Fig. 3A),** whereas SCZ perturbs excitatory neurons the most **(Fig. 3A)**. These patterns align with known biological insights related to these disorders. We also tracked the number of modules enriched with known risk genes and found the largest number of such modules in ASD **(Fig. 3A)**, likely due to relatively well-characterized knowledge bases for ASD.

**Figure 3:**
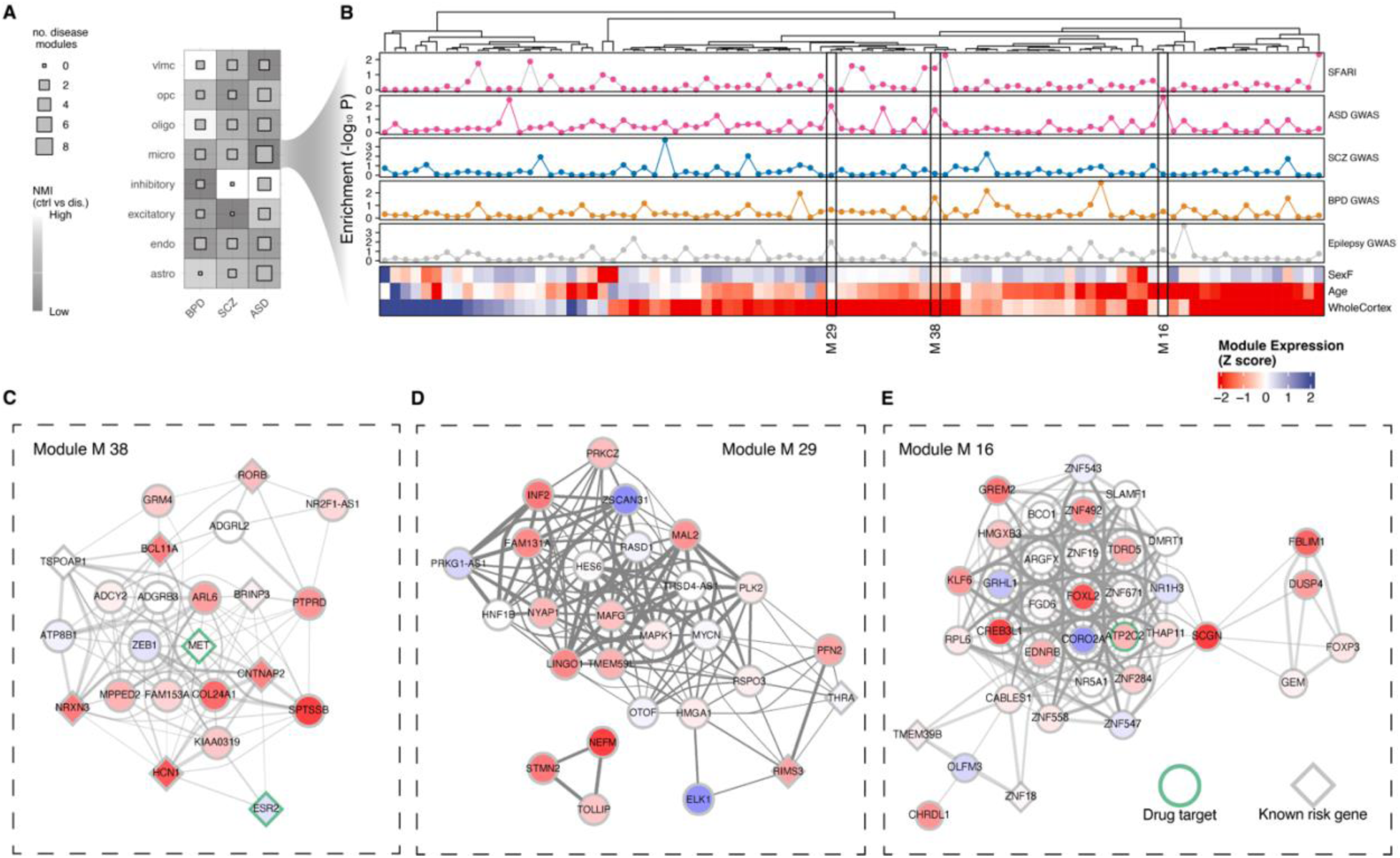
Prediction and analysis of co-regulated gene modules. **A)** Normalized Mutual Information (NMI) is used as a measure to gauge the deviation of network clustering between control and disorder networks. The heatmap shows the three disorders (x-axis), cell types (y-axis), and grids filled along a gray-to-white gradient representing low (dissimilar clustering) or high (similar clustering) mutual information, respectively. The inner squares are sized according to the number of disease modules identified within each cell type X disorder network. **B)** A zoom-out of microglial modules in ASD. The bottom-most panel of the heatmap shows modules (on the x-axis) and their differential expression (obtained from (*35*)) in sex, age, and whole cortex contrasts in ASD samples (y-axis). The grids of the heatmap are colored along a red-to-blue gradient indicating down- and- up-regulation, respectively. The columns of the heatmap are annotated for the enrichment of candidate ASD genes (from SFARI(*36*)) and GWAS-implicated loci in psychiatric disorders within the identified microglial modules (x-axis). Only three modules described in the main text are labeled. **C-E)** Visualization of modules M 38, M29, and M 16. Each circle represents a gene colored according to the log-fold change value in the whole cortex. Candidate risk genes are shaped as diamonds and the borders of drug targets are shaded green. Genes are connected by edges shaded along a gray gradient together with the thickness that is set to be proportional to the extent of regulator sharing.

Recent studies reveal shared genetic loci among these disorders, highlighting substantial overlap in their genetic underpinnings despite diagnostic and phenotypic complexity (*37*, *38*). Building on the, we further explore ASD modules to examine whether these modules are enriched for genetic variants (SNPs) associated with other psychiatric disorders. Using a heritability-aware enrichment analysis framework(*39*) and data from published GWAS studies, we identified several ASD microglial modules significantly enriched with SNPs linked to BPD, SCZ, and epilepsy **(Fig. 3B).** These findings suggest a convergence of genetic risk factors from multiple psychiatric disorders into common molecular modules, implying underlying shared molecular mechanisms, potentially underpinning overlapping clinical symptoms such as depression(*40*). One particular module of interest is module #38 (referred to as M38) which shows enrichment of both ASD GWAS, high-confidence gene sets from the SFARI database, as well as enrichment of bipolar GWAS (**Fig. 3B)**. Genes within M38 are collectively downregulated in whole cortex samples of ASD but remain relatively unchanged in age or sex (**Fig. 3B**). MET, a promising candidate gene for ASD(*41*, *42*), and a target for a BBB-permeable drug, is a member of M38 (**Fig. 3C**). Another interesting module with an incidence of hits in epilepsy and ASD GWAS is module M29 with down-regulated genes in whole coretex ASD (**Fig. 3B**) and membership of candidate ASD genes such as RIMS3(*43*) and THRA(*44*). We also recovered an age-specific ASD module, M16 (**Fig. 3B**), containing ASD candidate genes such as TMEM39B and ZNF18(*45*, *46*).

Collectively, our module analysis suggests that certain candidate risk genes are co-regulated in cell types. We find evidence of psychiatric disorders converging on a few of these modules, perhaps relevant to clinical phenotypes common across these disorders (e.g., depression), and possibly influenced by drug compounds with well-defined modes of action. This set of modules therefore provides a useful resource for more systematically analyzing drugs that may be repurposed to target specific cell clusters.

### Graph neural networks predict novel cell-type disorder genes

Most reported disease risk genes in the literature lack information on cell type specificity, which is a prerequisite for our main goal of cell type drug repurposing. Our analysis of network modules provides insight into genes that co-regulate with known candidate risk genes in certain cell types. Typically, a drug repurposing pipeline proceeds by analyzing a single disease module (identified based on enrichment of known risk genes) into a drug repurposing pipeline. However, such analyses rely heavily on the assumption that all genes within a disease module are equally equitable risk genes while leaving out several other risk genes that are distributed across several modules and vary based on the selected clustering algorithm. We address this limitation by using supervised graph convolutional networks (GCNs), training on cell-type-specific disorder gene regulatory networks (GRNs) where genes are represented as nodes and known risk genes, identified from public databases, are labeled. This allows us to leverage both known risk genes and cell-type networks as training data. Briefly, the GCN works as follows: A combination of graph convolution and fully connected layers are used to generate embeddings for each gene, predict directed edges between genes, and classify genes as risk or non-risk genes (**Fig. 4A; Supplementary Data S6**). The models are trained using 5-fold cross-validation. After validation, the average of all GCN disease scores was used for subsequent analyses.

**Figure 4:**
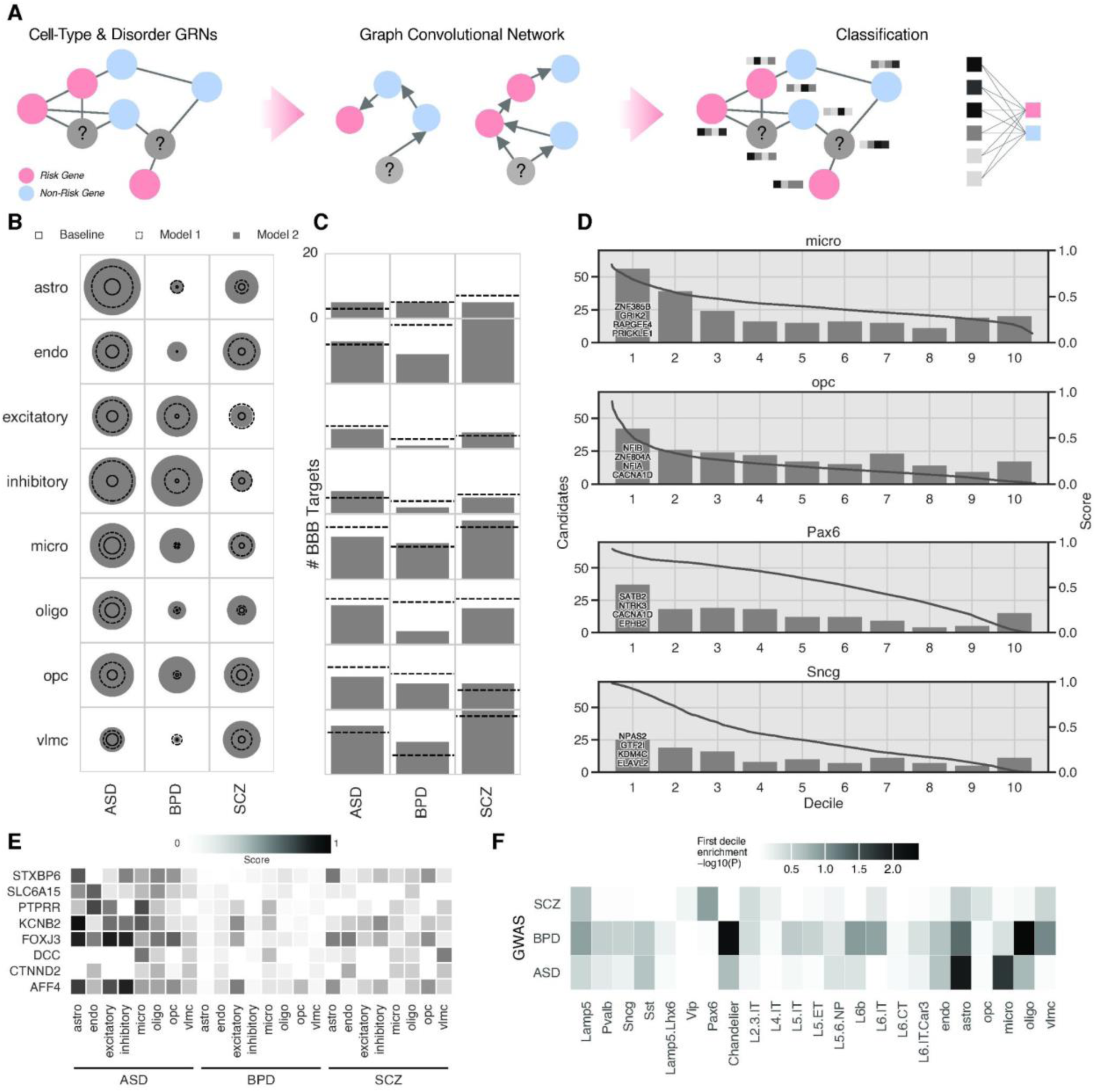
Graph learning model prioritizes existing candidate disease genes and novel risk genes in ASD, BPD, and SCZ. **A)** Schematic for gene classification using the graph learning model, i.e., graph convolutional network, featuring risk (red), non-risk (blue), and unknown (gray) genes. **B)** Performance of the model with specified targets. Circle sizes scale from radii of 0-1 based on AUPRC for baseline (solid line), model 1 (disease genes only, dashed line), and model 2 (disease and module genes, filled circle). **D)** Number of targets of blood-brain barrier (BBB) drugs found in the top decile of prioritized genes by model 1 (dashed line) and model 2 (filled bar) from 0 to 20. **D)** Histogram of the number of SFARI candidate category 2 genes found in each decile of ASD predicting models. The top decile is annotated with the top 4 prioritized candidate genes. The model score (right axis) is also plotted against the prediction percentile (gray line). **E)** Top 3 prioritized genes from each disease and their respective prioritizations within each cell type. Missing entries were not included in the corresponding cell type GRN. Cell types are categorized by disease and all scores are shaded based on per-model normalized risk scores. Darker squares indicate a higher model score. For panels B-E, results for additional cell types may be found in **Figure S7A-D**. **F)** Heatmap shows LD-aware enrichment of GWAS traits (y-axis) within the respective first decile predictions across cell types (x-axis). The grids are colored along a black gradient that is set to be proportional to the enrichment p-values, with darker colors indicating greater significance.

We train two models for each cell type based on different groups of risk genes. Model 1 is trained only on known disease genes, while model 2 incorporates genes from the modules discussed above. More details on the exact derivation of the gene lists can be found in **Supplementary Note 4.1**. We see that the AUPRC of model 2 is higher than that of model 1 in 21 of 24 cases for major cell types (**Fig. 4B**). Subclasses show a similar pattern, with 26 of 51 cell type-disease combinations exhibiting greater AUPRC for model 2 and 17 showing equal performance (**Fig. S7B**). To verify that this increased accuracy on modules does not come at the expense of biological relevance, we further examine the frequency of high prioritization for known targets of drugs that can cross the blood-brain barrier (BBB). Specifically, we compare the counts of BBB drug targets within the top decile of prioritized genes across models 1 and 2 (**Fig. 4C**). In general, the performance of models 1 and 2 is near-equivalent. Model 2 is favored in 11 cases, with 12 cases favoring model 1 out of a total of 24. This trend continues with subclasses, where 26 of 51 cases show equal performance between models 1 and 2 (**Fig. S7C**). Overall, model 2 achieves high AUPRC and prioritizes similar BBB drug targets compared to model 1. Going forward, model 2 is used for our analyses.

We have shown that incorporating genes from GRN modules improved the performance of our model, but it is also necessary to demonstrate the model’s reliability in predicting novel genes. We intentionally withheld SFARI candidate genes (category 2) from our ASD model to use for validation. When plotting candidate genes’ prioritizations, we observe a heavy skew toward the top decile (**Fig. 4D**). This skew is most pronounced in microglia and OPC, but is also seen in all major cell types aside from astrocytes and weakly in excitatory and inhibitory cell types except for Pax6 and Sncg (**Fig. S7C-D**). Our GCN is, therefore, able to effectively classify unseen risk genes with a high degree of reliability.

Finally, we predict novel genes using the trained models. Averaging gene prioritizations across all GRNs allows us to rank-order relevant genes and examine them for connections to ASD, BPD, and SCZ. In particular, we can take genes in the top decile of predictions for each cell type and disease and then pick only those genes that are present in all diseases (repeated for each cell type). This provides a list of 249 genes that were not positively labeled for one or more of the surveyed diseases (**Fig. 4E, Supplementary Data S6)**. For instance, CTNND2, a gene which encodes an adhesive junction associated protein of the armadillo/beta-catenin superfamily, is a listed category 2 gene in the SFARI genes database and was thus not used as a part of the training dataset. Yet, our model prioritizes CTNND2 for ASD. There is also more recent evidence that suggests the involvement of this gene in autism(*47*, *48*). Other examples include STXBP5, which has been noted as a seizure risk gene(*49*) with more recent evidence in ASD cases(*50*). SLC6A15 is additionally linked to major depressive disorder(*51*) and has been prioritized in ASD and BPD within overlapping gene sets for ASD, BPD, and SCZ(*52*). We also check the enrichment of genetic variants associated with the disorders in their respective GWAS studies within the top predictions. Interestingly, we find a greater enrichment of SNPs within non-neuronal cell types for ASD and BPD (**Fig. 4F**). Notably, BPD also showed significant enrichment in predictions for chandelier cells.

### Network-based drug repurposing

Network-based drug repurposing is an approach to use gene networks to bring existing molecules to new indications. This approach has been previously shown to be effective using cell type-naive protein interaction networks and cross-validated using EHR data(*23*). The unique dataset we have at hand presents an excellent opportunity to test the feasibility of drug repurposing to target specific cell clusters in psychiatric disorders. To do this, first, we assemble a BBB permeable drug-target network from public sources. This drug-target network has 802 drug molecules targeting 1192 genes (**Fig. S8; see Supplementary Note 5.1; Supplementary Data S7**). We observe an enrichment of known risk genes within this network, several of which are targeted by many drugs, especially GABA receptors **(Fig. S8).** We then leverage the cell type co-regulatory networks described above, together with predicted risk genes from the GNN model, for network-based drug repurposing using this drug-target network.

We calculate the minimum number of edges that need to be traversed to go from a set of drug targets set to a risk gene set (as predicted in our models) within a given cell type co-regulatory network. The hypothesis, as illustrated in **Figure 5A**, is that if the average path length between a drug target and a disease gene is short and non-random, the drug is likely to have a direct or close indirect effect on the disease pathway. We apply this procedure to all cell type networks we built across the three disorders. We find a total of 220 molecules that are significantly proximal (permutation-based tests, n=100; Z scores <= -3; **Supplementary Data S8**) to the predicted disorder risk genes. We find the largest distribution of repurposable drugs for SCZ and the least for ASD, which is counterintuitive given the stronger community confidence in known ASD risk genes in the literature. Overall, we find more repurposable drugs for deeper layer excitatory neurons (L6/IT, L6/CT, L6/IT/Car3), a few inhibitory neurons such as SST and LAMP5, and microglia **(Fig. S10A, B).** Collectively, a large fraction of the drugs that we found repurposable are neurotransmitter receptor modulators and reuptake inhibitors (**Fig. 5B**). For instance, the largest category of drugs belongs to serotonin reuptake inhibitors, which are widely used to treat depression.

**Figure 5:**
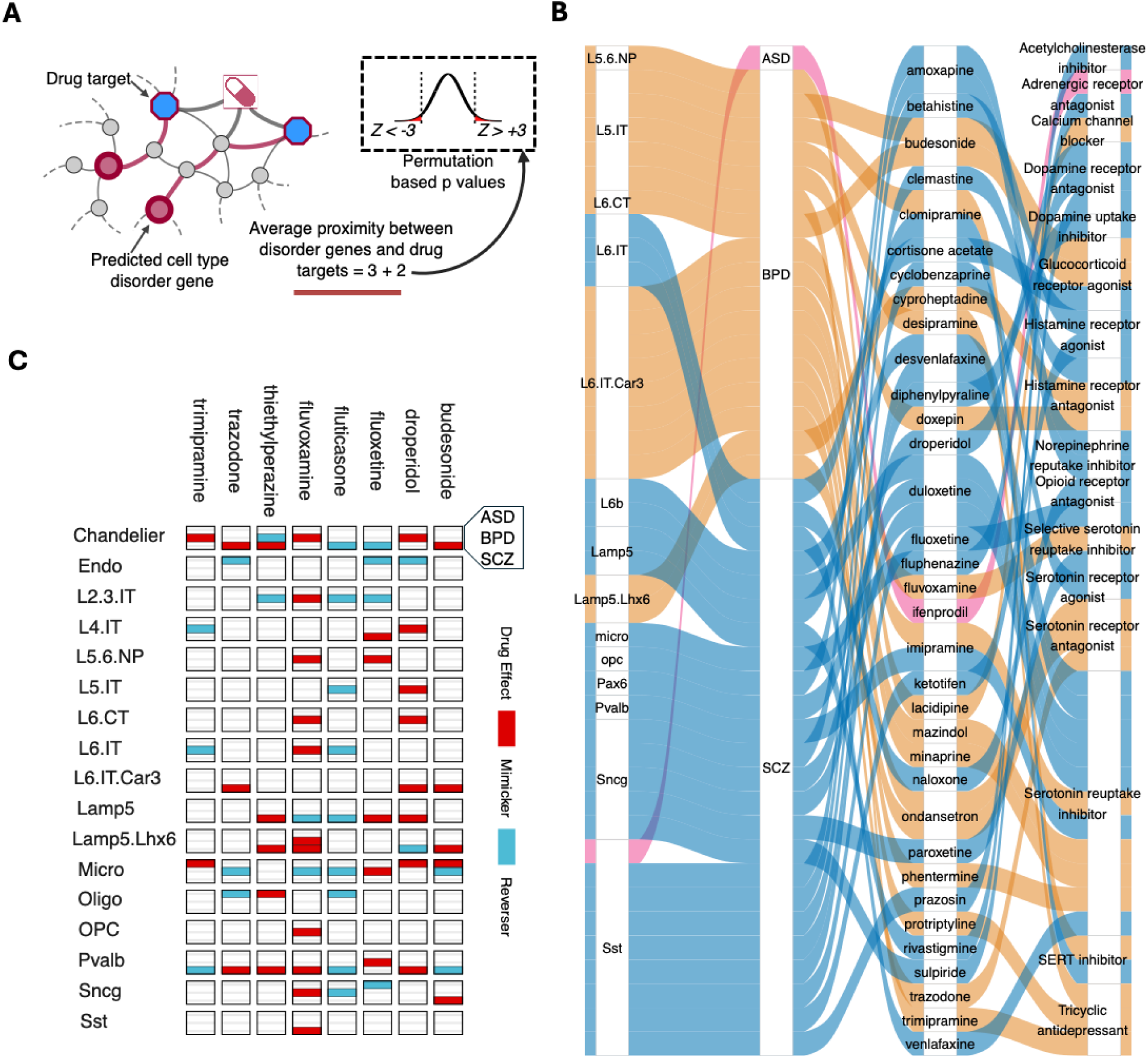
Network-based drug repurposing. **A)** An illustration depicting the network-based drug repurposing approach used in this study. Network proximity between cell type disorder genes (predicted in Figure 4) and listed drug targets are calculated across all cell type-disorder combinations. The observed proximity is then tested against a random background from which a Z score is derived. Repurposable drug candidates are chosen based on thresholding these Z scores. **B)** Sankey plot showing links between cell types (vertical axis 1) to disorders (vertical axis 2), drug molecules (vertical axis 3), and their mode of action (vertical axis 4) as predicted using the approach illustrated in **A**. **C)** The repurposable drug candidates are tested against gene expression data in the LINCS1000 dataset to determine whether the drug mimics or reverses expression of gene sets observed under disorder conditions in the PEC2 dataset(*53*) (**see Supplementary Note 5.3**). The heatmap shows cell types and disorders on the y-axis and a collection of top drug molecules (based on Z scores) on the x-axis. The grids are divided into three rows, one for each disorder, and colored red for mimickers and blue for reversers.

To verify whether the drugs nominated by our approach can modulate the transcriptional phenotype of cells, we used the gene expression profiles from the Integrated Network-based Cellular Signatures (LINCS) project(*54*). The LINCS L1000 dataset provides gene expression measurements from various human cell lines that have been treated with thousands of small molecules. By comparing the direction of gene expression changes induced by disorders with those caused by drugs, we identified 37 drugs in our nominated drug list that mimic or reverse the gene expression patterns caused by disorders in a cell type dependent manner (**Fig. 5C; Supplementary Data S9**). Specifically, mimickers are drugs that replicate the gene expression changes observed in the disorder, aligning in the same direction. In contrast, reversers are drugs that counteract these changes, shifting gene expression in the opposite direction, which suggests a potential to mitigate or correct the molecular disruptions caused by the disorder. For example, genes that are differentially expressed upon fluoxetine treatment (a selective serotonin reuptake inhibitor (SSRI) that is used to treat depression, OCD, and BPD) show opposite expression patterns of those observed in chandelier cells from SCZ donors, endothelial cells from ASD donors, and L2/3 IT neurons from BPD donors (**Fig. 5C**). Fluvoxamine, another SSRI, shows potential for targeting SCZ-perturbed Lamp5 cells and BPD-perturbed microglial cells (**Fig. 5C**).

We accurately recover a number of drugs, such as asenapine, an atypical antipsychotic used for the treatment of SCZ and acute mania associated with BPD. We find that asenapine has close proximity to SCZ risk genes, mostly for neuronal cell types (**Fig. S10C**). Another interesting example of a correctly recovered drug for SCZ is Iloperidone, an antipsychotic that is already used to treat SCZ symptoms. Our data suggest that Iloperidone reverses gene expression, especially in several neuronal cells (**Fig. S10D**).

### Drug-associated cell-type eQTLs

Pharmacogenetics is a well-developed field focused on how different patients exhibit disparate responses to therapeutic compounds, and how these patient-specific differences may be driven by differences in genotype. For instance, one critical mechanism by which genotypic differences may influence differences in drug response is one in which genotypic variation drives changes in the expression of drug target genes. As a demonstration of this idea, a very simple example scenario may be one in which an eQTL manifests as a variant that is associated with substantially diminished expression of a gene targeted by a particular drug. Under this simple scenario, patients the harbor that variant may fail to respond to therapy, given that the drug would effectively have no target gene on which to act. Critically, these general types of mechanisms are likely to play out at the level of distinct cell types.

The catalog of BBB drug targets used in our study (combined with genotypic as well as cell type-level expression data) provide uniquely powerful building blocks to explore pharmacogenetic phenomena of this nature in greater depth than previously possible. As different therapeutic compounds affect different sets of genes, we sought to evaluate the extent to which genotypic variation is associated with significant changes in expression for the gene sets that are targeted by different drugs. To do so, we formulate a “drug-associated cell-type eQTL” analysis by measuring the extent to which expression variation of drug targets is associated with variants throughout the genome. One initial motivation for this analysis was the observation that up to 17% overlap between drug targets and genes in our recently reported cell type eQTLs(*8*, *55–57*) (**Fig. 6A**), which indicates that a substantial proportion of druggable gene targets exhibit expression that is strongly associated with genetic variation in specific cell types.

**Figure 6.**
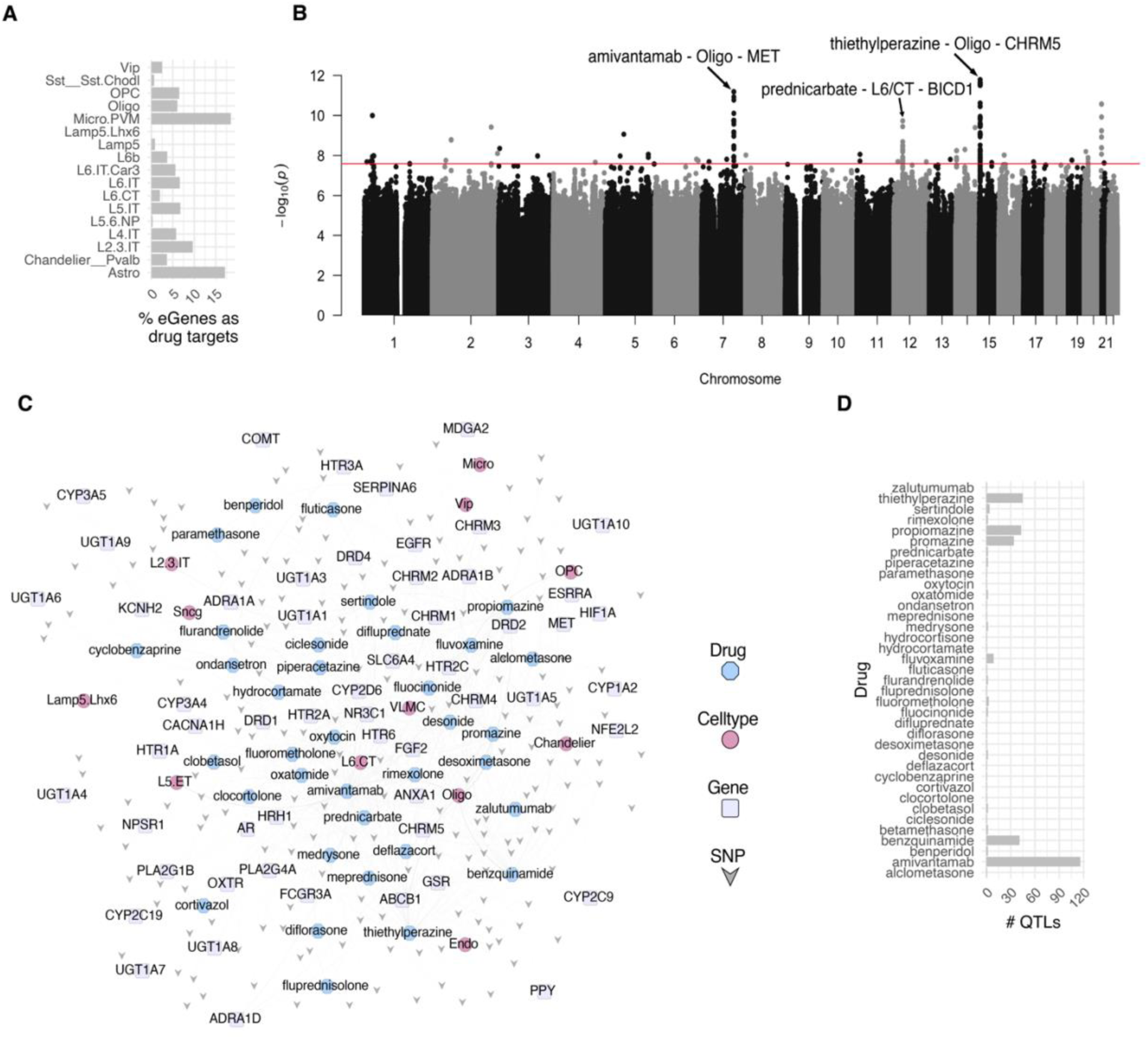
Drug-associated cell-type eQTLs. **A)** Fraction of the number of cell type cis-eQTL eGenes (as reported in (*8*)) that are also listed targets of BBB-permeable drugs. **B)** Manhattan plot showing p-values (y-axis) and variants’ genomic coordinates (x-axis) from the drug-associated cell-type eQTL analysis. The analysis was performed using expression values of drug targets for those drugs that were found to be repurposable in our analysis. **C)** Statistically significant drug-associated cell-type eSNPs (shaped ‘V’) are plotted alongside associated drugs (octagons), their target genes (round rectangles), and associated cell types (circles) as a network graph. **D)** Distribution of the number of SNPs (x-axis) associated with drug QTLs (y-axis).

For each drug in each cell type, we use its set of target genes to build a single composite gene expression value by simply calculating the mean log-normalized expression of a given drug’s targets within each cell type. Thus, instead of using the expression of a single gene as the phenotype, a given phenotype is given as a composite gene expression score for a drug-cell type pair. Under this framework, we identify 335 eQTLs, which are defined as significant drug-associated cell type eQTLs (**Supplementary Data S10)**. Here, significance is determined using a Bonferroni-corrected p-value threshold of 0.05, with Bonferroni correction being applied at the level of each phenotype – that is, for a given drug-cell type combination, Bonferroni correction was applied by adjusting for ∼2 million tests, where ∼2 million is the number of variants tested in our search for QTLs (further details are given in **Supplementary Note 6)**. These eQTLs may be interpreted as cases wherein a drug-associated gene set collectively exhibits strong associations with genotypic variation. As such, they highlight cases for which responses to drug exposure may vary considerably across samples, with this variation being strongly influenced by genotype. The results from this analysis are summarized in **Fig. 6B**.

We visualized statistically significant eSNPs along with the connected drugs, target genes, and associated cell types as a network which reveals 12 of the 23 cell types we included in our analysis, with oligodendrocytes a prominent hub (**Fig. 6C**). Interestingly, we found the largest number of variants linked to propiomazine and promazine, two antipsychotics used to treat positive and negative symptoms of SCZ (**Fig. 6D**). This suggests that genetic variation between individuals may influence responses to these specific drugs, potentially offering insight into variability in drug efficacy. Interestingly, we also found other potentially repurposable drugs that are not currently used to treat psychiatric disorders. For instance, Benzquinamide, a hub in our network, is an antihistamine with sedative properties. Benzquinamide’s ability to antagonize dopamine and histamine receptors suggests that it could have some potential impact on psychiatric symptoms. Histamine H1 antagonism can lead to sedation, which might be useful for patients with agitation or insomnia, which frequently accompany psychiatric disorders. However, these claims remain subject to intensive clinical trials, which is beyond the scope of our study.

## Discussion

In this study, we used a population-level single-cell approach to integrate principles of network medicine with analyses focused on elucidating the genetic basis of psychiatric disorders. Our approach allowed us to investigate several molecular and therapeutic aspects of ASD, BPD, and SCZ. First, our approach expands the druggable genome by focusing on transcription factor (TF) regulons within specific cell types. Our analysis identifies key regulons enriched with differentially expressed genes that are also listed targets of drug molecules that are predicted to cross the blood-brain barrier. Second, using a combination of unsupervised (co-regulated gene modules) and supervised (graph neural network) machine learning approaches, we identified novel genes associated with psychiatric disorders. These genes act as a lever for a more nuanced analysis of repurposable drugs at the cell type level, a strategy previously not possible due to a lack of sufficient datasets with replicate cohorts. Third, our network-based drug repurposing strategy nominates several FDA-approved drug molecules, some of which can be independently verified for their potential to reverse the transcriptional phenotypes of disorders in brain cell types. Fourth, our exploration of DrugCell QTLs provides crucial insights into the intrinsic expression variability of drug targets, thereby offering a foundation for developing personalized treatment strategies based on an individual’s genetic profile.

The GRN resource and accompanying analyses provide insights into dysregulated gene expression in psychiatric disorders, both within and between cell types **(Fig. 2B, S4A-C)**. For instance, the GRNs inform those drugs that can reverse the expression of certain TFs that are differentially expressed in diagnosed brains **(Fig. 2E-F)**. Such TFs provide a handle on regulatory switches as novel targets for drug discovery. For instance, PBX1, a high-confidence ASD candidate(*45*), is downregulated in ASD, a pattern that may be reversed by certain drugs **(Fig. S4D).** Furthermore, the GRNs are useful in generating modules of co-regulated genes that can be aligned with any independent single nuclear or bulk RNA-seq dataset (in a framework much like that of GO enrichment) to identify modules relevant to the experiment **(Fig. 3B)**. We found certain modules that harbor genes implicated across multiple psychiatric and related disorders. These modules provide a handle on targets that can be leveraged to develop drugs that span multiple disorders with common clinical phenotypes in psychiatry (e.g. depression). Furthermore, the GRNs allow us to train machine learning models to predict novel risk genes. For each cell type, we provide a list of such genes across the three disorders.

Our study details an approach for expanding network-based drug discovery, previously limited to cell-type-naive PPI networks, to single-cell genomics within cellular contexts. This allows us to identify drugs that can target specific cell types. For instance, we found a large number of drugs for excitatory neurons in BPD, whereas inhibitory neurons seem to be more amenable to treating SCZ **(Fig. 5B)**. The prioritization of a large number of serotonin uptake inhibitors, typically used to treat depression, indicates that our approach mostly identifies compounds that are designed to treat symptoms common across psychiatric disorders. This also suggests the development of more refined models that can pinpoint drug combinations that better suit the peculiarities of individual disorders. It should also be noted that although we cross-verified the expression patterns of genes differentially expressed in disorder with those in response to perturbations by a few drugs, extensive experimental validation in human brain cell lines remains a limitation of our study. Furthermore, a large fraction of the LINCS L1000 dataset we used is imputed and limited to neuronal cells from the CNS. Thus, the reverser and mimicker effects we identify would need to be further refined with observations in multiple cell lines of the human brain. Note that the repurposable drugs nominated by our study are simply computational predictions and would thus need thorough clinical validation in follow-up studies.

The drug-associated cell type eQTLs approach allowed us to identify genomic variants that exhibit strong associations with variability in the expression of drug target genes. To the extent that patient-specific drug response is influenced by the underlying expression of a given drug’s target genes, these drug-associated cell type eQTLs identify specific genomic loci that may influence drug response in genotype-specific ways by modulating expression at the level of distinct cell types. Thus, by integrating genetic variation and drug targeting, our analysis provides a framework to better understand how genotype can impact drug response at the cellular level, thereby offering new insights into precision medicine-based strategies. However, we note that our framework should not be interpreted as offering drug response QTLs in a direct sense, as that would entail using drug response data from the same group of individuals for which the genotypes are available, which currently remains limited in publicly available datasets. Nevertheless, the drug-Cell QTLs identified can potentially offer valuable guides for patient stratification in clinical trials. For instance, our results list drugs that might be more susceptible to response variation among individuals (drugs with a large number of associated SNPs).

## Data Availability

The GRNs generated in this study, along with repurposable drugs, can be downloaded from Zenodo repository. Additionally, these data can be parsed using a web-app hosted at https://daifengwanglab.shinyapps.io/psyDGN/.

## Materials and Methods Summary

The Materials and Methods for each section of the Main Text are available in the Supplementary Materials, *which are organized using the same section headings as in the main text*. These include a detailed description of the data subsets assessed in the network medicine analysis and all computational and statistical analyses performed for each part of the analysis.

## Acknowledgments

We would like to express our deep gratitude to the patients and their families who generously donated the invaluable biological material essential for the success of this study. We are profoundly indebted to their participation and commitment to advancing scientific knowledge and improving human health.

## Funding

We acknowledge the National Institutes of Health grants, RF1MH128695 (to D.W.), U24MH136793 (to M.G.), R21NS127432 (to D.W.), R21NS128761 (to D.W.), U01MH116492 (to D.W.), P50HD105353 (to Waisman Center), National Science Foundation Career Award 2144475 (to D.W.), Simons Foundation Autism Research Initiative pilot grant 971316 (to D.W.), and the start-up funding for D.W. from the Office of the Vice Chancellor for Research and Graduate Education at the University of Wisconsin–Madison. The funders had no role in study design, data collection and analysis, decision to publish, or manuscript preparation

## Author contributions

All individually named authors contributed substantially to the paper through either data generation or analysis: data generation: K.H.A., S.K., X.Z., Y.Z.; and data analysis: C.G., N.C.K., D.C., J.C. The following two corresponding authors co-led the analysis: D.W. and M.G.

## Competing interests

The authors declare no competing interests.

## Supplementary materials

Materials and Methods

Figs. S1 to S10

Data S1 to S10

## Notes

### Competing Interest Statement

The authors have declared no competing interest.

## References

1. Y. Agid, G. Buzsáki, D. M. Diamond, R. Frackowiak, J. Giedd, J.-A. Girault, A. Grace, J. J. Lambert, H. Manji, H. Mayberg, M. Popoli, A. Prochiantz, G. Richter-Levin, P. Somogyi, M. Spedding, P. Svenningsson, D. Weinberger, How can drug discovery for psychiatric disorders be improved? Nat Rev Drug Discov 6, 189–201 (2007).

2. N. G. Skene, J. Bryois, T. E. Bakken, G. Breen, J. J. Crowley, H. A. Gaspar, P. Giusti-Rodriguez, R. D. Hodge, J. A. Miller, A. B. Muñoz-Manchado, M. C. O’Donovan, M. J. Owen, A. F. Pardiñas, J. Ryge, J. T. R. Walters, S. Linnarsson, E. S. Lein, Major Depressive Disorder Working Group of the Psychiatric Genomics Consortium, P. F. Sullivan, J. Hjerling-Leffler, Genetic identification of brain cell types underlying schizophrenia. Nat Genet 50, 825–833 (2018).

3. B. Wamsley, L. Bicks, Y. Cheng, R. Kawaguchi, D. Quintero, J. Grundman, J. Liu, S. Xiao, N. Hawken, M. Margolis, S. Mazariegos, D. H. Geschwind, Molecular cascades and cell-type specific signatures in ASD revealed by single cell genomics. bioRxiv, 2023.03.10.530869 (2023).

4. W. B. Ruzicka, S. Mohammadi, J. F. Fullard, J. Davila-Velderrain, S. Subburaju, D. R. Tso, M. Hourihan, S. Jiang, H.-C. Lee, J. Bendl, PsychENCODE Consortium§, G. Voloudakis, V. Haroutunian, G. E. Hoffman, P. Roussos, M. Kellis, PsychENCODE Consortium, Single-cell multi-cohort dissection of the schizophrenia transcriptome. Science 384, eadg5136 (2024).

5. H. Mathys, J. Davila-Velderrain, Z. Peng, F. Gao, S. Mohammadi, J. Z. Young, M. Menon, L. He, F. Abdurrob, X. Jiang, A. J. Martorell, R. M. Ransohoff, B. P. Hafler, D. A. Bennett, M. Kellis, L.-H. Tsai, Single-cell transcriptomic analysis of Alzheimer’s disease. Nature 570, 332–337 (2019).

6. K. E. Prater, K. J. Green, S. Mamde, W. Sun, A. Cochoit, C. L. Smith, K. L. Chiou, L. Heath, S. E. Rose, J. Wiley, C. D. Keene, R. Y. Kwon, N. Snyder-Mackler, E. E. Blue, B. Logsdon, J. E. Young, A. Shojaie, G. A. Garden, S. Jayadev, Human microglia show unique transcriptional changes in Alzheimer’s disease. Nat Aging 3, 894–907 (2023).

7. S. G. Lago, J. Tomasik, G. F. van Rees, H. Steeb, D. A. Cox, N. Rustogi, J. M. Ramsey, J. Bishop, T. Petryshen, S. J. Haggarty, J. Vázquez-Bourgon, S. Papiol, P. Suarez-Pinilla, B. Crespo-Facorro, N. J. van Beveren, S. Bahn, Drug discovery for psychiatric disorders using high-content single-cell screening of signaling network responses ex vivo. Sci Adv 5, eaau9093 (2019).

8. P. S. Emani, J. J. Liu, D. Clarke, M. Jensen, J. Warrell, C. Gupta, R. Meng, C. Y. Lee, S. Xu, C. Dursun, S. Lou, Y. Chen, Z. Chu, T. Galeev, A. Hwang, Y. Li, P. Ni, X. Zhou, PsychENCODE Consortium‡, T. E. Bakken, J. Bendl, L. Bicks, T. Chatterjee, L. Cheng, Y. Cheng, Y. Dai, Z. Duan, M. Flaherty, J. F. Fullard, M. Gancz, D. Garrido-Martín, S. Gaynor-Gillett, J. Grundman, N. Hawken, E. Henry, G. E. Hoffman, A. Huang, Y. Jiang, T. Jin, N. L. Jorstad, R. Kawaguchi, S. Khullar, J. Liu, J. Liu, S. Liu, S. Ma, M. Margolis, S. Mazariegos, J. Moore, J. R. Moran, E. Nguyen, N. Phalke, M. Pjanic, H. Pratt, D. Quintero, A. S. Rajagopalan, T. R. Riesenmy, N. Shedd, M. Shi, M. Spector, R. Terwilliger, K. J. Travaglini, B. Wamsley, G. Wang, Y. Xia, S. Xiao, A. C. Yang, S. Zheng, M. J. Gandal, D. Lee, E. S. Lein, P. Roussos, N. Sestan, Z. Weng, K. P. White, H. Won, M. J. Girgenti, J. Zhang, D. Wang, D. Geschwind, M. Gerstein, PsychENCODE Consortium, Single-cell genomics and regulatory networks for 388 human brains. Science 384, eadi5199 (2024).

9. J. E. Darnell, Transcription factors as targets for cancer therapy. Nat Rev Cancer 2, 740– 749 (2002).

10. J. Liu, N. B. Perumal, C. J. Oldfield, E. W. Su, V. N. Uversky, A. K. Dunker, Intrinsic disorder in transcription factors. Biochemistry 45, 6873–6888 (2006).

11. J. H. Bushweller, Targeting transcription factors in cancer - from undruggable to reality. Nat Rev Cancer 19, 611–624 (2019).

12. J. E. Darnell, Transcription factors as targets for cancer therapy. Nat Rev Cancer 2, 740– 749 (2002).

13. P. Wu, Q. Feng, V. E. Kerchberger, S. D. Nelson, Q. Chen, B. Li, T. L. Edwards, N. J. Cox, E. J. Phillips, C. M. Stein, D. M. Roden, J. C. Denny, W.-Q. Wei, Integrating gene expression and clinical data to identify drug repurposing candidates for hyperlipidemia and hypertension. Nat Commun 13, 46 (2022).

14. S. de Jong, L. R. Vidler, Y. Mokrab, D. A. Collier, G. Breen, Gene-set analysis based on the pharmacological profiles of drugs to identify repurposing opportunities in schizophrenia. J Psychopharmacol 30, 826–830 (2016).

15. H.-C. So, C. K.-L. Chau, A. Lau, S.-Y. Wong, K. Zhao, Translating GWAS findings into therapies for depression and anxiety disorders: gene-set analyses reveal enrichment of psychiatric drug classes and implications for drug repositioning. Psychol Med 49, 2692– 2708 (2019).

16. P. Csermely, V. Agoston, S. Pongor, The efficiency of multi-target drugs: the network approach might help drug design. Trends Pharmacol Sci 26, 178–182 (2005).

17. W. Sun, P. E. Sanderson, W. Zheng, Drug combination therapy increases successful drug repositioning. Drug Discov Today 21, 1189–1195 (2016).

18 I. F. N. Hung, K. K. W. To, J. F. W. Chan, V. C. C. Cheng, K. S. H. Liu, A. Tam, T.-C. Chan, J. Zhang, P. Li, T.-L. Wong, R. Zhang, M. K. S. Cheung, W. Leung, J. Y. N. Lau, M. Fok, H. Chen, K.-H. Chan, K.-Y. Yuen, Efficacy of Clarithromycin-Naproxen-Oseltamivir Combination in the Treatment of Patients Hospitalized for Influenza A(H3N2) Infection: An Open-label Randomized, Controlled, Phase IIb/III Trial. Chest 151, 1069–1080 (2017).

19. L. Wang, Y. Lu, D. Li, Y. Zhou, L. Yu, I. Mesa Eguiagaray, H. Campbell, X. Li, E. Theodoratou, The landscape of the methodology in drug repurposing using human genomic data: a systematic review. Brief Bioinform 25, bbad527 (2024).

20. T. T. T. Truong, Z. S. J. Liu, B. Panizzutti, J. H. Kim, O. M. Dean, M. Berk, K. Walder, Network-based drug repurposing for schizophrenia. Neuropsychopharmacology 49, 983– 992 (2024).

21. Z. Mortezaei, J.-B. Cazier, A. A. Mehrabi, C. Cheng, A. Masoudi-Nejad, Novel putative drugs and key initiating genes for neurodegenerative disease determined using network-based genetic integrative analysis. J Cell Biochem 120, 5459–5471 (2019).

22. C. Gupta, J. Xu, T. Jin, S. Khullar, X. Liu, S. Alatkar, F. Cheng, D. Wang, Single-cell network biology characterizes cell type gene regulation for drug repurposing and phenotype prediction in Alzheimer’s disease. PLoS Comput Biol 18, e1010287 (2022).

23. J. Fang, P. Zhang, Y. Zhou, C.-W. Chiang, J. Tan, Y. Hou, S. Stauffer, L. Li, A. A. Pieper, J. Cummings, F. Cheng, Endophenotype-based in silico network medicine discovery combined with insurance record data mining identifies sildenafil as a candidate drug for Alzheimer’s disease. Nat Aging 1, 1175–1188 (2021).

24. D. Morselli Gysi, Í. do Valle, M. Zitnik, A. Ameli, X. Gan, O. Varol, S. D. Ghiassian, J. J. Patten, R. A. Davey, J. Loscalzo, A.-L. Barabási, Network medicine framework for identifying drug-repurposing opportunities for COVID-19. Proc Natl Acad Sci U S A 118, e2025581118 (2021).

25. N. Selvaraj, A. K. Swaroop, B. S. S. Nidamanuri, R. R. Kumar, J. Natarajan, J. Selvaraj, Network-based Drug Repurposing: A Critical Review. Curr Drug Res Rev 14, 116–131 (2022).

26. F. Cheng, R. J. Desai, D. E. Handy, R. Wang, S. Schneeweiss, A.-L. Barabási, J. Loscalzo, Network-based approach to prediction and population-based validation of in silico drug repurposing. Nat Commun 9, 2691 (2018).

27. P. Paci, G. Fiscon, F. Conte, R.-S. Wang, L. Farina, J. Loscalzo, Gene co-expression in the interactome: moving from correlation toward causation via an integrated approach to disease module discovery. NPJ Syst Biol Appl 7, 3 (2021).

28. R.-T. Hu, Q. Yu, S.-D. Zhou, Y.-X. Yin, R.-G. Hu, H.-P. Lu, B.-L. Hu, Co-expression Network Analysis Reveals Novel Genes Underlying Alzheimer’s Disease Pathogenesis. Front Aging Neurosci 12, 605961 (2020).

29. S. van Dam, U. Võsa, A. van der Graaf, L. Franke, J. P. de Magalhães, Gene co-expression analysis for functional classification and gene-disease predictions. Brief Bioinform 19, 575– 592 (2018).

30. N. Safari-Alighiarloo, M. Taghizadeh, M. Rezaei-Tavirani, B. Goliaei, A. A. Peyvandi, Protein-protein interaction networks (PPI) and complex diseases. Gastroenterol Hepatol Bed Bench 7, 17–31 (2014).

31. H. Wang, J. Yang, Y. Zhang, J. Wang, Discover novel disease-associated genes based on regulatory networks of long-range chromatin interactions. Methods 189, 22–33 (2021).

32. L. Fan, J. Hou, G. Qin, Prediction of Disease Genes Based on Stage-Specific Gene Regulatory Networks in Breast Cancer. Front Genet 12, 717557 (2021).

33. P. Buphamalai, T. Kokotovic, V. Nagy, J. Menche, Network analysis reveals rare disease signatures across multiple levels of biological organization. Nat Commun 12, 6306 (2021).

34. J.-J. Zhuang, Q. Liu, D.-L. Wu, L. Tie, Current strategies and progress for targeting the “undruggable” transcription factors. Acta Pharmacol Sin 43, 2474–2481 (2022).

35. M. J. Gandal, J. R. Haney, B. Wamsley, C. X. Yap, S. Parhami, P. S. Emani, N. Chang, G. T. Chen, G. D. Hoftman, D. de Alba, G. Ramaswami, C. L. Hartl, A. Bhattacharya, C. Luo, T. Jin, D. Wang, R. Kawaguchi, D. Quintero, J. Ou, Y. E. Wu, N. N. Parikshak, V. Swarup, T. G. Belgard, M. Gerstein, B. Pasaniuc, D. H. Geschwind, Broad transcriptomic dysregulation occurs across the cerebral cortex in ASD. Nature 611, 532–539 (2022).

36. B. S. Abrahams, D. E. Arking, D. B. Campbell, H. C. Mefford, E. M. Morrow, L. A. Weiss, I. Menashe, T. Wadkins, S. Banerjee-Basu, A. Packer, SFARI Gene 2.0: a community-driven knowledgebase for the autism spectrum disorders (ASDs). Mol Autism 4, 36 (2013).

37. L. S. Carroll, M. J. Owen, Genetic overlap between autism, schizophrenia and bipolar disorder. Genome Med 1, 102 (2009).

38. J. E. Rexach, Y. Cheng, L. Chen, D. Polioudakis, L.-C. Lin, V. Mitri, A. Elkins, X. Han, M. Yamakawa, A. Yin, D. Calini, R. Kawaguchi, J. Ou, J. Huang, C. Williams, J. Robinson, S. E. Gaus, S. Spina, E. B. Lee, L. T. Grinberg, H. Vinters, J. Q. Trojanowski, W. W. Seeley, D. Malhotra, D. H. Geschwind, Cross-disorder and disease-specific pathways in dementia revealed by single-cell genomics. Cell 187, 5753–5774.e28 (2024).

39. C. A. de Leeuw, J. M. Mooij, T. Heskes, D. Posthuma, MAGMA: generalized gene-set analysis of GWAS data. PLoS Comput Biol 11, e1004219 (2015).

40. N. Craddock, M. J. Owen, The Kraepelinian dichotomy - going, going… but still not gone. Br J Psychiatry 196, 92–95 (2010).

41. D. B. Campbell, R. D’Oronzio, K. Garbett, P. J. Ebert, K. Mirnics, P. Levitt, A. M. Persico, Disruption of cerebral cortex MET signaling in autism spectrum disorder. Ann Neurol 62, 243–250 (2007).

42. I. Thanseem, K. Nakamura, T. Miyachi, T. Toyota, S. Yamada, M. Tsujii, K. J. Tsuchiya, A. Anitha, Y. Iwayama, K. Yamada, E. Hattori, H. Matsuzaki, K. Matsumoto, Y. Iwata, K. Suzuki, S. Suda, M. Kawai, G.-I. Sugihara, K. Takebayashi, N. Takei, H. Ichikawa, T. Sugiyama, T. Yoshikawa, N. Mori, Further evidence for the role of MET in autism susceptibility. Neurosci Res 68, 137–141 (2010).

43. R. A. Kumar, J. Sudi, T. D. Babatz, C. W. Brune, D. Oswald, M. Yen, N. J. Nowak, E. H. Cook, S. L. Christian, W. B. Dobyns, A de novo 1p34.2 microdeletion identifies the synaptic vesicle gene RIMS3 as a novel candidate for autism. J Med Genet 47, 81–90 (2010).

44. M. Pilhatsch, C. Winter, K. Nordström, B. Vennström, M. Bauer, G. Juckel, Increased depressive behaviour in mice harboring the mutant thyroid hormone receptor alpha 1. Behav Brain Res 214, 187–192 (2010).

45. S. De Rubeis, X. He, A. P. Goldberg, C. S. Poultney, K. Samocha, A. E. Cicek, Y. Kou, L. Liu, M. Fromer, S. Walker, T. Singh, L. Klei, J. Kosmicki, F. Shih-Chen, B. Aleksic, M. Biscaldi, P. F. Bolton, J. M. Brownfeld, J. Cai, N. G. Campbell, A. Carracedo, M. H. Chahrour, A. G. Chiocchetti, H. Coon, E. L. Crawford, S. R. Curran, G. Dawson, E. Duketis, B. A. Fernandez, L. Gallagher, E. Geller, S. J. Guter, R. S. Hill, J. Ionita-Laza, P. Jimenz Gonzalez, H. Kilpinen, S. M. Klauck, A. Kolevzon, I. Lee, I. Lei, J. Lei, T. Lehtimäki, C.-F. Lin, A. Ma’ayan, C. R. Marshall, A. L. McInnes, B. Neale, M. J. Owen, N. Ozaki, M. Parellada, J. R. Parr, S. Purcell, K. Puura, D. Rajagopalan, K. Rehnström, A. Reichenberg, A. Sabo, M. Sachse, S. J. Sanders, C. Schafer, M. Schulte-Rüther, D. Skuse, C. Stevens, P. Szatmari, K. Tammimies, O. Valladares, A. Voran, W. Li-San, L. A. Weiss, A. J. Willsey, T. W. Yu, R. K. C. Yuen, DDD Study, Homozygosity Mapping Collaborative for Autism, UK10K Consortium, E. H. Cook, C. M. Freitag, M. Gill, C. M. Hultman, T. Lehner, A. Palotie, G. D. Schellenberg, P. Sklar, M. W. State, J. S. Sutcliffe, C. A. Walsh, S. W. Scherer, M. E. Zwick, J. C. Barett, D. J. Cutler, K. Roeder, B. Devlin, M. J. Daly, J. D. Buxbaum, Synaptic, transcriptional and chromatin genes disrupted in autism. Nature 515, 209–215 (2014).

46. M. H. Chahrour, T. W. Yu, E. T. Lim, B. Ataman, M. E. Coulter, R. S. Hill, C. R. Stevens, C. R. Schubert, ARRA Autism Sequencing Collaboration, M. E. Greenberg, S. B. Gabriel, C. A. Walsh, Whole-exome sequencing and homozygosity analysis implicate depolarization-regulated neuronal genes in autism. PLoS Genet 8, e1002635 (2012).

47. A. J. S. Chan, W. Engchuan, M. S. Reuter, Z. Wang, B. Thiruvahindrapuram, B. Trost, T. Nalpathamkalam, C. Negrijn, S. Lamoureux, G. Pellecchia, R. V. Patel, W. W. L. Sung, J. R. MacDonald, J. L. Howe, J. Vorstman, N. Sondheimer, N. Takahashi, J. H. Miles, E. Anagnostou, K. Tammimies, M. Zarrei, D. Merico, D. J. Stavropoulos, R. K. C. Yuen, B. A. Fernandez, S. W. Scherer, Genome-wide rare variant score associates with morphological subtypes of autism spectrum disorder. Nat Commun 13, 6463 (2022).

48. H. Mendez-Vazquez, R. L. Roach, K. Nip, S. Chanda, M. F. Sathler, T. Garver, R. A. Danzman, M. C. Moseley, J. P. Roberts, O. N. Koch, A. A. Steger, R. Lee, J. Arikkath, S. Kim, The autism-associated loss of δ-catenin functions disrupts social behavior. Proc Natl Acad Sci U S A 120, e2300773120 (2023).

49. H. N. Cukier, N. D. Dueker, S. H. Slifer, J. M. Lee, P. L. Whitehead, E. Lalanne, N. Leyva, I. Konidari, R. C. Gentry, W. F. Hulme, D. V. Booven, V. Mayo, N. K. Hofmann, M. A. Schmidt, E. R. Martin, J. L. Haines, M. L. Cuccaro, J. R. Gilbert, M. A. Pericak-Vance, Exome sequencing of extended families with autism reveals genes shared across neurodevelopmental and neuropsychiatric disorders. Mol Autism 5, 1 (2014).

50. X. Zhou, P. Feliciano, C. Shu, T. Wang, I. Astrovskaya, J. B. Hall, J. U. Obiajulu, J. R. Wright, S. C. Murali, S. X. Xu, L. Brueggeman, T. R. Thomas, O. Marchenko, C. Fleisch, S. D. Barns, L. G. Snyder, B. Han, T. S. Chang, T. N. Turner, W. T. Harvey, A. Nishida, B. J. O’Roak, D. H. Geschwind, SPARK Consortium, J. J. Michaelson, N. Volfovsky, E. E. Eichler, Y. Shen, W. K. Chung, Integrating de novo and inherited variants in 42,607 autism cases identifies mutations in new moderate-risk genes. Nat Genet 54, 1305–1319 (2022).

51. M. A. Kohli, S. Lucae, P. G. Saemann, M. V. Schmidt, A. Demirkan, K. Hek, D. Czamara, M. Alexander, D. Salyakina, S. Ripke, D. Hoehn, M. Specht, A. Menke, J. Hennings, A. Heck, C. Wolf, M. Ising, S. Schreiber, M. Czisch, M. B. Müller, M. Uhr, T. Bettecken, A. Becker, J. Schramm, M. Rietschel, W. Maier, B. Bradley, K. J. Ressler, M. M. Nöthen, S. Cichon, I. W. Craig, G. Breen, C. M. Lewis, A. Hofman, H. Tiemeier, C. M. van Duijn, F. Holsboer, B. Müller-Myhsok, E. B. Binder, The neuronal transporter gene SLC6A15 confers risk to major depression. Neuron 70, 252–265 (2011).

52. J. Guan, J. J. Cai, G. Ji, P. C. Sham, Commonality in dysregulated expression of gene sets in cortical brains of individuals with autism, schizophrenia, and bipolar disorder. Transl Psychiatry 9, 152 (2019).

53. P. S. Emani, J. J. Liu, D. Clarke, M. Jensen, J. Warrell, C. Gupta, R. Meng, C. Y. Lee, S. Xu, C. Dursun, S. Lou, Y. Chen, Z. Chu, T. Galeev, A. Hwang, Y. Li, P. Ni, X. Zhou, PsychENCODE Consortium, T. E. Bakken, J. Bendl, L. Bicks, T. Chatterjee, L. Cheng, Y. Cheng, Y. Dai, Z. Duan, M. Flaherty, J. F. Fullard, M. Gancz, D. Garrido-Martín, S. Gaynor-Gillett, J. Grundman, N. Hawken, E. Henry, G. E. Hoffman, A. Huang, Y. Jiang, T. Jin, N. L. Jorstad, R. Kawaguchi, S. Khullar, J. Liu, J. Liu, S. Liu, S. Ma, M. Margolis, S. Mazariegos, J. Moore, J. R. Moran, E. Nguyen, N. Phalke, M. Pjanic, H. Pratt, D. Quintero, A. S. Rajagopalan, T. R. Riesenmy, N. Shedd, M. Shi, M. Spector, R. Terwilliger, K. J. Travaglini, B. Wamsley, G. Wang, Y. Xia, S. Xiao, A. C. Yang, S. Zheng, M. J. Gandal, D. Lee, E. S. Lein, P. Roussos, N. Sestan, Z. Weng, K. P. White, H. Won, M. J. Girgenti, J. Zhang, D. Wang, D. Geschwind, M. Gerstein, Single-cell genomics and regulatory networks for 388 human brains. bioRxiv, 2024.03.18.585576 (2024).

54. J. E. Evangelista, D. J. B. Clarke, Z. Xie, A. Lachmann, M. Jeon, K. Chen, K. M. Jagodnik, S. L. Jenkins, M. V. Kuleshov, M. L. Wojciechowicz, S. C. Schürer, M. Medvedovic, A. Ma’ayan, SigCom LINCS: data and metadata search engine for a million gene expression signatures. Nucleic Acids Res 50, W697–W709 (2022).

55. A. K. Malhotra, G. M. Murphy, J. L. Kennedy, Pharmacogenetics of psychotropic drug response. Am J Psychiatry 161, 780–796 (2004).

56. M. Pirmohamed, Pharmacogenomics: current status and future perspectives. Nat Rev Genet 24, 350–362 (2023).

57. M. R. Nelson, T. Johnson, L. Warren, A. R. Hughes, S. L. Chissoe, C.-F. Xu, D. M. Waterworth, The genetics of drug efficacy: opportunities and challenges. Nat Rev Genet 17, 197–206 (2016).

