## Supplementary Materials for "Network-based drug repurposing for psychiatric disorders using single-cell genomics"

#### Materials and Methods

##### 1 Cell-type gene regulatory network construction for drug repurposing in psychiatric disorders

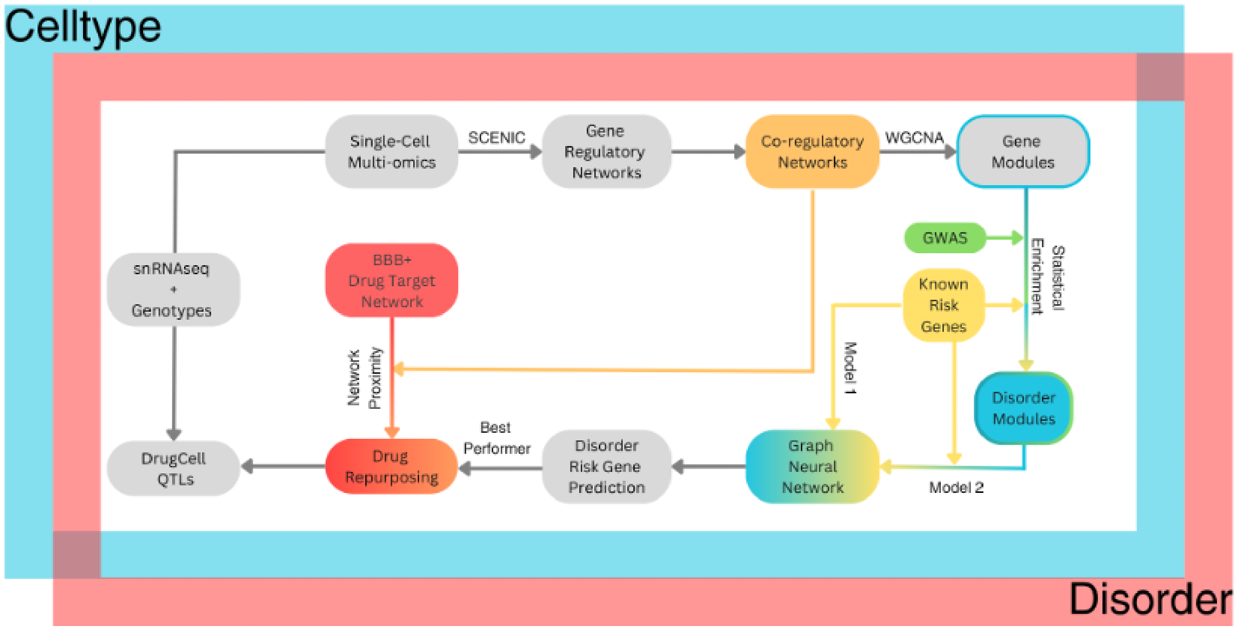


**Figure S1: Single-Cell Network-Medicine workflow.** The flowchart depicts the flow of data for network-based drug repurposing. The directed GRNs predicted by integrating snRNA-seq and scATAC-seq were transformed into coregulatory networks on the basis of overlaps between predicted regulators of every pair of target genes. The coregulatory networks were then clustered to find modules of highly coregulated genes. The identified modules were than investigated for enrichment of GWAS SNPs and known risk genes across each disorder. Subsequently, two GNN models were trained to identify and prioritize risk genes at the cell type level across disorders. The first model used known cell type naive risk genes and the second model used known cell type naive risk genes plus genes within the top cell type module enriched with risk genes or GWAS. The best performing model was chosen to then select prioritized cell type disorder genes to be subsequently used for drug repurposing. The drugs identified as repurposable were then subjected to Drug-associated cell-type eQTL analysis which utilized snRNA-seq and genotype data.

#### 1.1 snRNA-seq and scATAC-seq data processing

The psychENCODE2 (PEC2) dataset encompassing snRNA-seq and scATAC-seq samples from 388 donors has been uniformly processed as described in the recent publication(*1*). We used the pre-processed data, including cell clustering and annotation schemes used in the original publication, for the analysis presented here. Figure S1 depicts the flow of data through our network-based drug repurposing pipeline (**Fig. S1**). We created two sets of disorder GRNs for each cell type. The first set of GRNs are built for only major cell type clusters, namely, excitatory and inhibitory neurons, astrocytes, oligodendrocytes, oligodendrocyte progenitor cells (OPC), microglia, vlmc, and endothelial cells. Furthermore, for each disorder, we build control and disorder GRNs for each cell type separately. This split of the dataset allowed us to make cross-cohort contrasts and comparisons, as well as test for reproducibility for disorders with redundant cohorts. We built the second set of GRNs for subclass level cell types which include 'L2.3.IT', 'L4.IT', 'L5.IT', 'L5.ET', 'L5.6.NP', 'L6b', 'L6.IT', 'L6.CT', and 'L6.IT.Car3' subclasses of excitatory neurons, and 'Lamp5', 'Pvalb', 'Sncg', 'Sst', 'Lamp5.Lhx6', 'Vip', 'Pax6', and ’Chandelier’ subclasses of inhibitory neurons. The set 2 cell type GRNs were built for each disorder separately, combining redundant cohorts for SCZ. Altogether, cells from a total of 247 donors (36% females) were used for GRN inference. The data subset comprises a total of 1,64,5813 cells, with 901,101 (54.76%) disorder cells and 744,712 (45.24%) control cells (**Fig. S2**). The following procedure was applied to predict the cell type by disorder GRNs.

#### 1.2 Gene Regulatory Networks

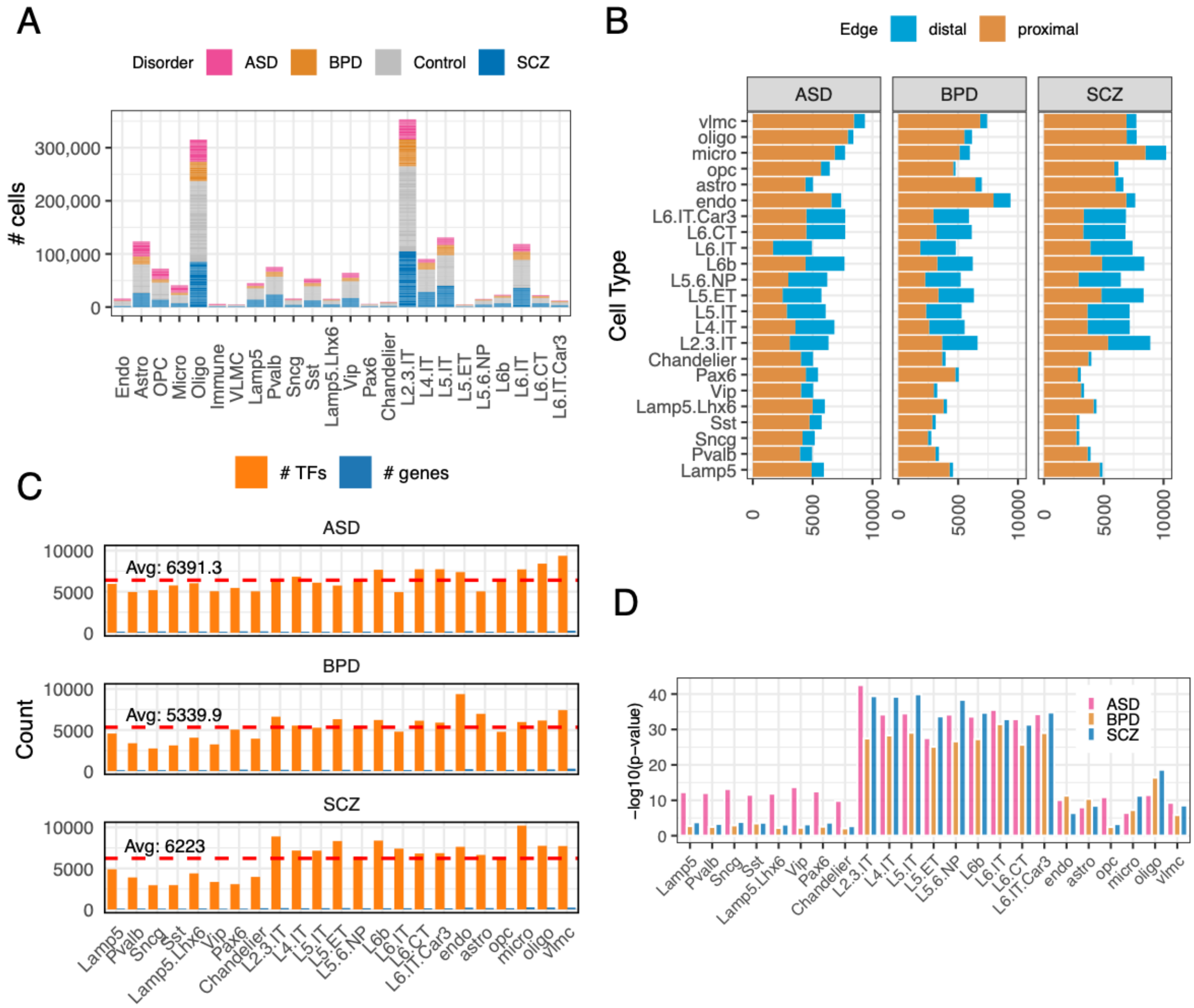


**Figure S2. Summary of cell type disorder GRNs.** A) Number of cells in snRNA-seq data (y-axis) used for GRN inference across cell types (x-axis). The bars are stacked and colored uniquely for each disorder and controls. B) Distribution of proximal and distal edges (TF→TG links; x-axis) across cell types (y-axis) and disorders (facets). C) Distribution of the number of TFs and target genes (y-axis) across cell types (x-axis). The red dotted line depicts the average number of genes across cell types for the particular disorder. D) Enrichment of enhancer-regulated drug targets represented as -log10 (p value) (y-axis) across cell types (x-axis) with bars colored uniquely for each disorder.

We first normalized raw counts within a cell cluster using the *'NormalizeData'* function of Seurat v5(*2*). We then filtered genes with low variation in expression across a cell cluster in a given cohort using Seurat's *'FindVariableFeatures'*(*3*) and created a list of top 5000 highly variable genes (HVG). To the HVG list, we added TFs for which binding sites (TFBS) are known(*4*). We reason that because TFs are regulatory genes, subtle changes in their expression patterns can have profound downstream effects. Excluding such TFs from analysis may collapse the GRN making it sparse. Subsequently, we supplied the expression matrix consisting of appended HVGs to the GRNboost2 algorithm(*5*), along with the list of TFs, to predict their targets.

Subsequently, we provided the output of GRNBoost2 to SCENIC to filter TF→TG links that are not representative of TFBS within gene promoters(*6*). Briefly, SCENIC uses cisTarget to perform motif enrichment analysis by scanning the promoters of coexpressed genes to identify overrepresented TF binding motifs. Only transcription factors whose motifs are statistically enriched are considered potential regulators of the genes. Such TF→TG links were labeled as proximal edges. For distal links, we used enhancer-promoter links from scATAC-seq data and mapped TFBS on these interaction regions, thereby linking TFs to targets via enhancers (as described previously in the original PEC2 publication(*7*)). We further filtered the links from scATAC-seq data if there was no coexpression between the TF→TG pair in the cell cluster (detected in GRNboost2 runs) and labeled the remaining links as distal edges. This step allowed us to add variation to distal links in sub class GRNs since the scATAC-seq data is only available for the major class cell types. Finally, for each cell type disorder combination, we merged the proximal and distal edges into composite GRNs made available for further analyses.

#### 1.3 GRN evaluation

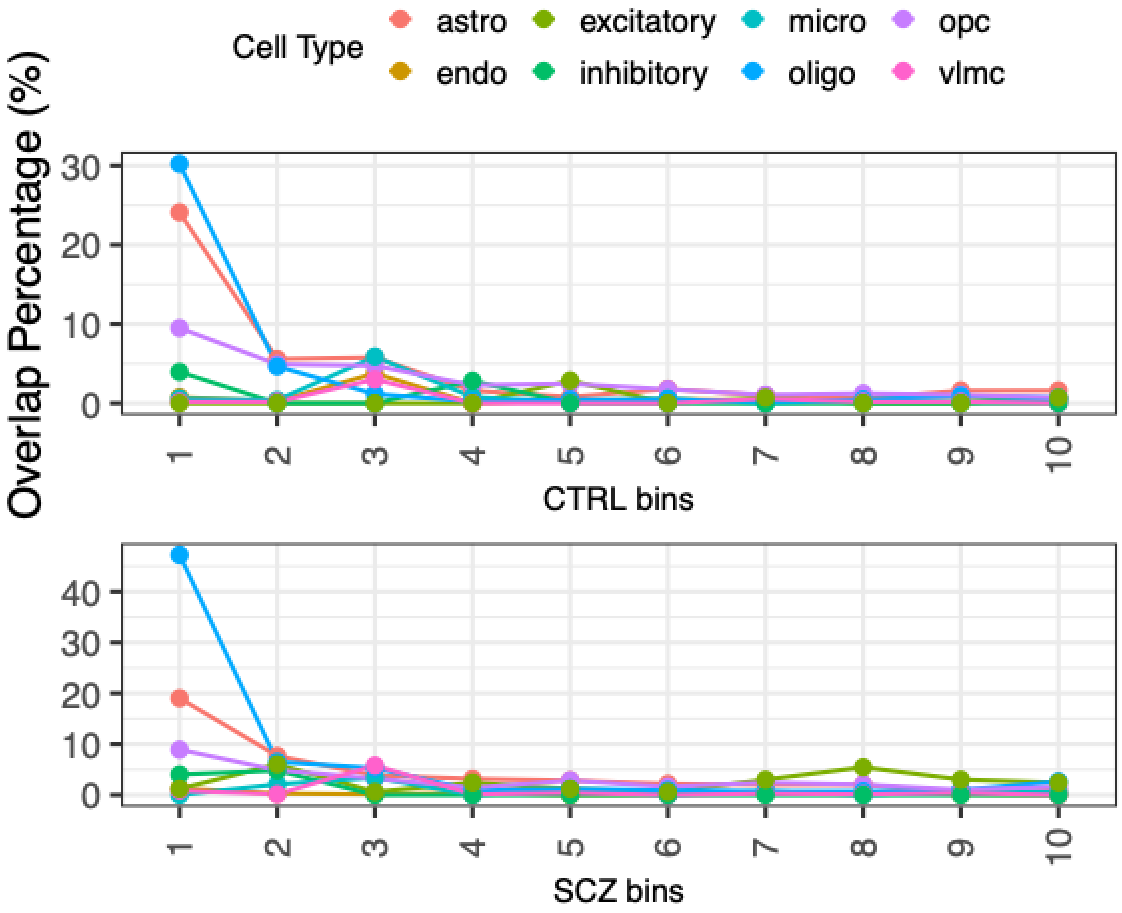


**Figure S3. Evaluation of cross-cohort GRNs.** A) The plot shows the overlaps between proximal edges in control (top) and schizophrenia (SCZ; bottom) within two independent cohorts. The y-axis shows the overlap percent within each of the 10 bins created based on the decreasing order of edge importance scores.

To assess the robustness of the inferred GRNs, we examined the sensitivity of our pipeline to sampling biases. We grouped the TF→TG links into ten deciles based on decreasing edge importance scores as inferred by SCENIC. We then calculated the percentage of edge overlap in each decile between SCZ GRNs from the CMC and SZBD cohorts for each cell type, repeating the process for control GRNs as well. Our analysis revealed a relatively higher overlap in the top deciles, with a gradual decline and minimal overlap in the 10th decile (**Fig. S3**). This suggests that high-confidence GRN links are reproducible, while lower-confidence links may be more sensitive to sampling variations.

### 2 Druggable cell-type transcription factors for psychiatric disorders

#### 2.1 TF centrality analysis and disease influence scores

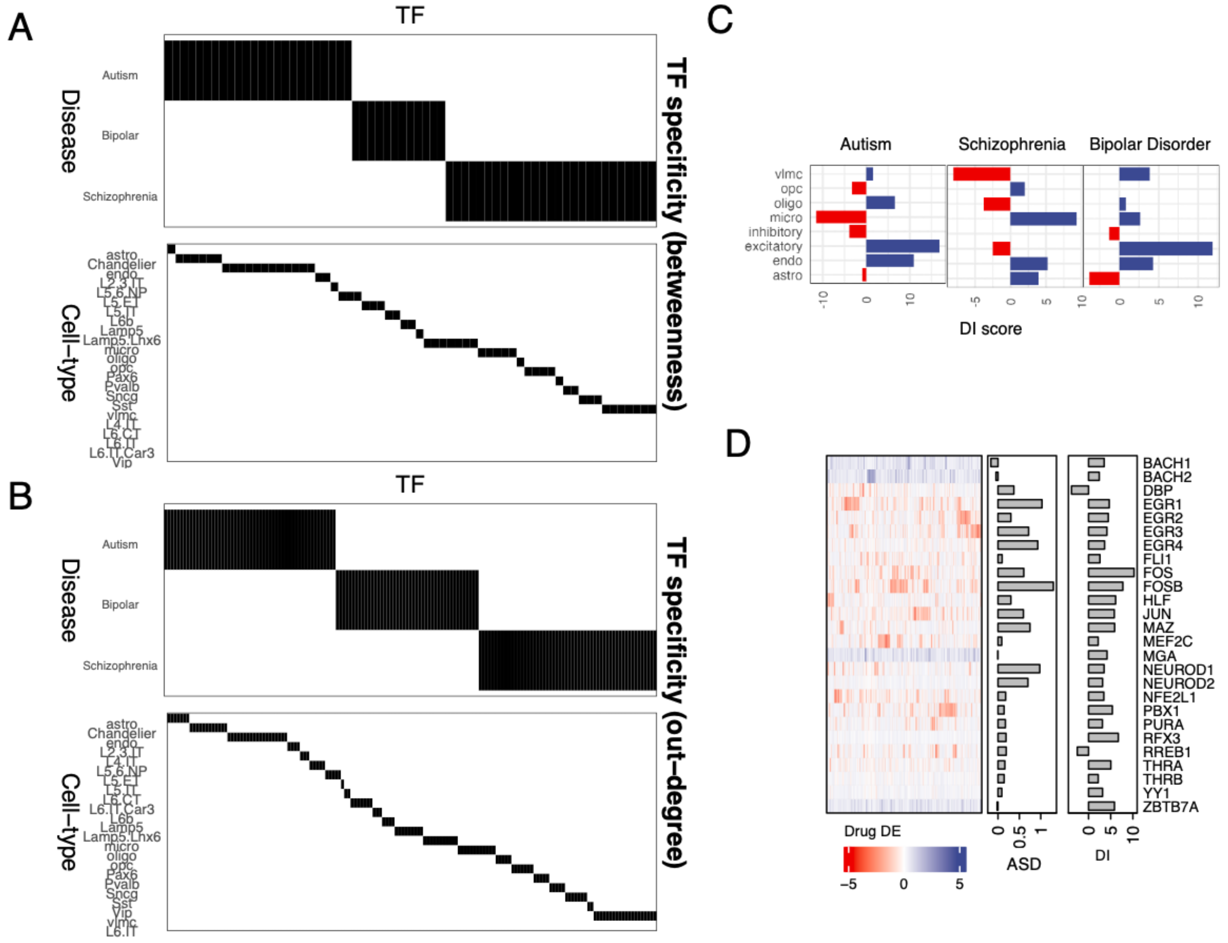


**Figure S4. Centrality analysis of GRNs.** A-B) TFs that are specific to cell types and disorders were mined using betweenness centrality (A) and outdegrees (B). TFs are listed along the x-axis and disorders on the top y-axis and celltypes on the bottom y-axis. The girds of the heatmap are shaded black if the TF is found in the top 10% list of the corresponding cell type and disorder GRN, white if not. C) Cumulative

DI scores of TFs within individual cell types across the three disorders. D) TFs with the largest changes in DI scores are shown on the y-axis along with their DI scores (right barplot) and log of change in expression in disorder w.r.t. control (left barplot). The heatmaps show change in expression of TFs in CNS cells treated with drug compounds (x-axis; names hidden) represented along a blue to red gradient indicating upregulation or downregulation, respectively.

#### 2.2 Hubs, bottlenecks, and disorder influence score

We counted the number of outgoing edges from a given TF in a given GRN as its outdegree. We used betweenness centrality to quantify the number of times a TF appears in the shortest paths between pairs of target genes. To assess the influence of TFs in disease contexts, we calculated the Disease Influence (DI) score using betweenness centrality as a proxy. The DI score of a TF is defined as the log2 ratio of the betweenness centrality in disease gene regulatory networks (GRNs) to the betweenness centrality in control GRNs (**Fig. S5**).

$$\mathrm{DI} = \log_2\left(\frac{BC_{\mathrm{disease}}}{BC_{\mathrm{control}}}\right)$$

Where $BC_{\mathrm{disease}}\ and\ BC_{\mathrm{control}}$ are betweenness centrality in disease and control GRNs, respectively.

#### 2.3 TF specificities

We profiled TF specificity using betweenness centrality and outdegree for diseases and cell types. We selected TFs based on their top 10% outdegrees and betweenness centralities, respectively. We then counted the occurrence of these TFs across diseases and cell types and filtered out TFs that appeared in only one disease or cell type (referred to as disease-specific TFs and cell type-specific TFs, respectively). Finally, we identified TFs that are specific to diseases and to cell types based on outdegree, and separately identified TFs that are specific to diseases and to cell types based on betweenness centrality. We categorized these TFs into disease-specific and cell type-specific groups based on betweenness centrality and outdegree, respectively (**Fig. S4 A,B**).

#### 2.4 Druggable Regulons

To identify druggable regulons, we obtained gene expression profiles from chemically perturbed human cell lines available through the SigCom LINCS website(*8*). Specifically, we worked with the Characteristic Direction (CD)-processed L1000 signatures. The CD method is a computational approach used to analyze gene expression data within the Library of Integrated Network-based Cellular Signatures (LINCS) L1000 dataset. Its primary objective is to pinpoint the most significant gene expression changes in response to various perturbations. The CD method claims to offer a robust ranking of differentially expressed genes, based on their contribution to the overall perturbation effect(*9*).

Next, we obtained a list of drugs predicted to permeate the blood-brain barrier (BBB) from the B3DB database(*10*) (see Section 5.1 for further details) and intersected these BBB-permeable drugs with those in the LINCS L1000 CD dataset. For the matching drugs, we compared each TF's expression in response to chemical perturbations in the LINCS dataset to the gene expression observed in the respective disorders. We found several TFs with high DI scores whose expression can be reversed by certain drugs, as shown in **Figure 2 E-F and S4D.**

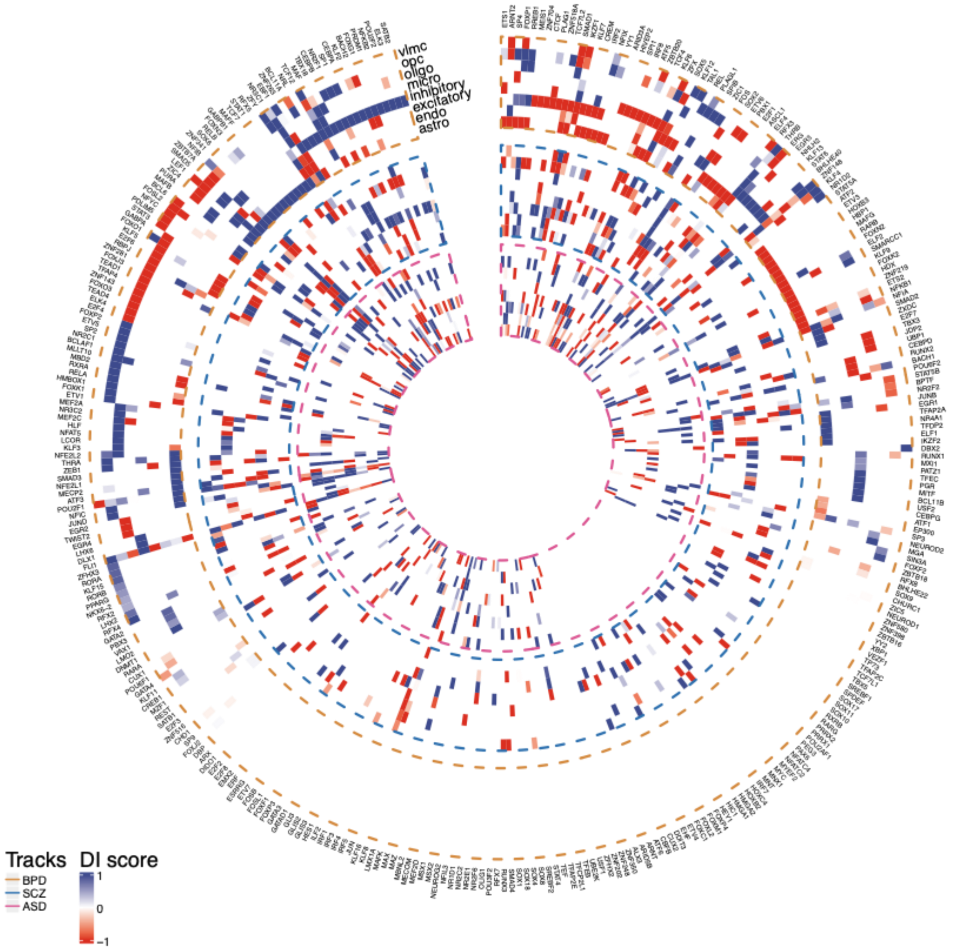


**Figure S5. Disease influence scores of TFs.** The circular heatmap shows distribution of TF disease influence (DI) scores across cell types and disorders. The outermost, middle, and inner rings show distributions in bipolar  disorder (BPD), schizophrenia (SCZ), and autism (ASD), respectively. TFs with are labeled on the y-axis. The cells of the heatmap are colored along a blue to red gradient representing gain or loss in influence, respectively. Only the outer ring of the heatmap is clustered, the inner two rings remain unclustered to preserve order.

### 3 Disorder risk genes converge on coregulated gene modules

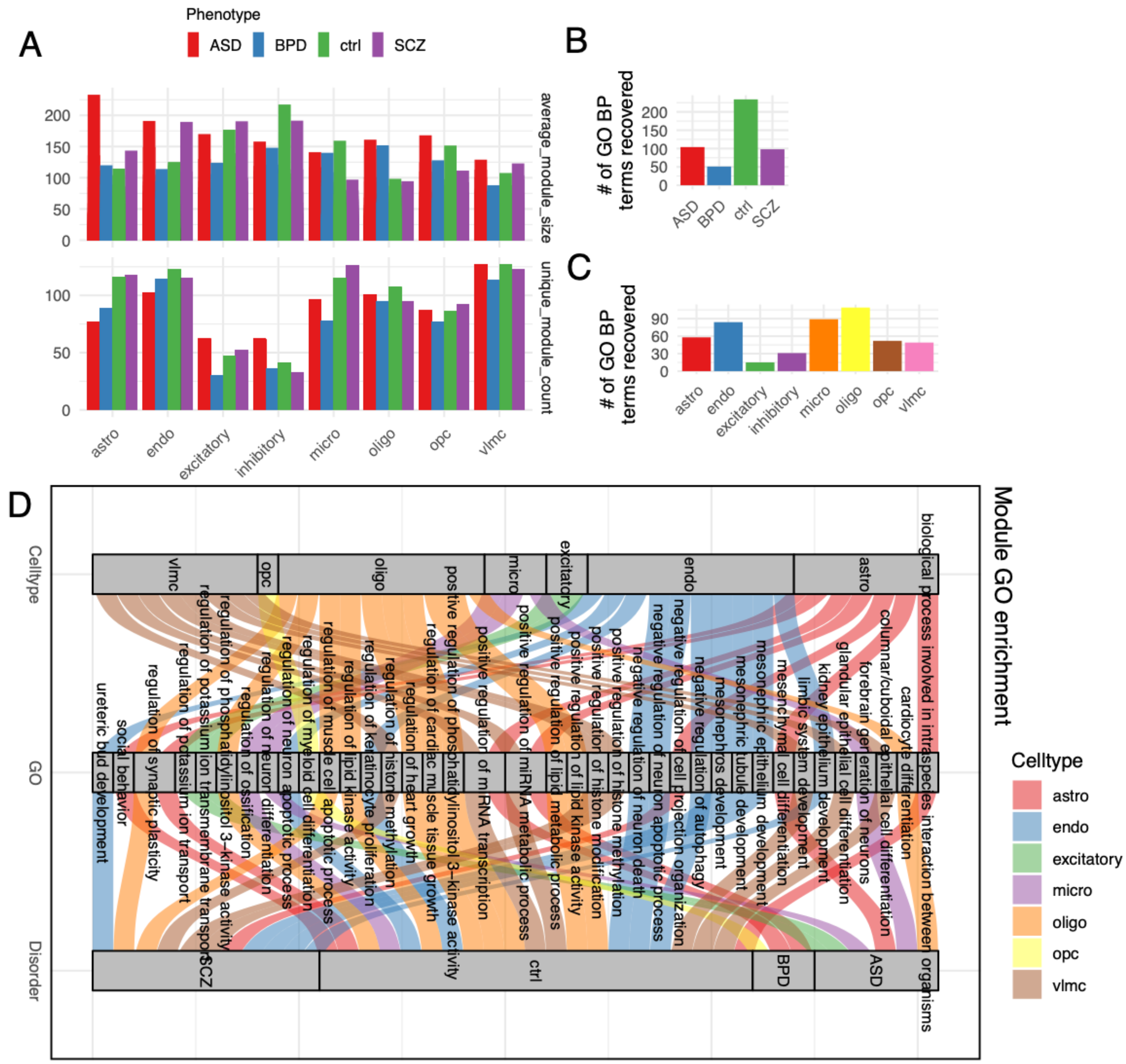


**Figure S6. Coregulated gene modules.** A) The top barplot shows average size (y-axis) of modules identified across cell types (x-axis). The lower plot shows number of modules (y-axis) identified within each cell type (x-axis). The bars are colored uniquely for each disorder as shown in the legend above. B-C) The number of GO BP terms found significantly enriched (hypergeometric tests corrected p-values <= 0.1) within each disorder across all cell types (B) and within each cell type across all disorders (C). D) A sankey plot showing GO BP terms (middle) enriched within cell type modules (axis 1) and disorders (axis 3).

#### 3.1 Coregulation modules

Our main goal was to prioritize repurposable drugs for the three psychiatric disorders in a cell-type specific manner using a network based approach. In our initial survey, we found only one TF (PPARG) listed as a direct drug target. This indicated that most drug targets are non-TF genes. Therefore, it is imperative to GRNs from the perspective of non-TF, or target genes (TG). To align with the GRNs presented in this study, we converted the TF→ TG edges into TG→ TG edges using the following procedure.

First, we used the Jaccard's Index (JI) to quantify the overlap between the regulators of a TG pair in a GRN. We then arranged the resulting JI values as a dissimilarity matrix (1 - JI) with TGs in rows and columns. The dissimilarity matrix was then supplied to a hierarchical clustering algorithm using the *flashClust* function of WGCNA(*11*). The dendrograms of the resulting clustering were then cut using *cutreeDynamic* function (with parameters *method="hybrid", deepSplit = 2, pamRespectsDendro = FALSE, msize = 30*). The *msize = 30* parameter ensures a minimum module size of 30 for a respectable sample size for downstream enrichment analysis as described below. Note that we used only proximal links for identification of modules because the shared distal links between control and disorder might obscure biological signals.

#### 3.2 Magma enrichment

We used the MAGMA (Multi-marker Analysis of GenoMic Annotation) to perform LD aware enrichment of GWAS traits within module gene sets and other gene sets described in the main text. To do this, first we obtained a number of GWAS results and extracted SNPs with nominal p-values ($p <= 0.05$). The SNPs were then annotated by applying the *annotate* function using gene locations from the human genome build 38 obtained from NCBI. The reference files used for enrichment were based on European ancestry in Phase 3 of the 1,000 Genomes project. The sample size parameter *N* was set according to the sample size mentioned in the corresponding GWAS publications as follows: SCZ: 76755(*12*), ASD: 46350(*13*), BPD: 41917(*14*), epilepsy: 29944(*15*). The resulting p values from MAGMA runs are shown in the main **Figure 3B**.

#### 3.3 Gene Ontology enrichment

To test the enrichment of gene sets from the GO catalog(*16*), we downloaded the GMT formatted GO term annotations from the Enrichment Map database available (https://download.baderlab.org/EM_Genesets/). The GMT files were read into the R using the GSA library (version 1.03.2). Then, biological process terms with more than 300 or less than 30 genes were filtered and the remaining terms used to calculate the over-representation of annotated genes annotated within each of the query gene sets (e.g. modules). The statistical significance of the overlap was calculated using a hypergeometric test using the sum of all genes across all modules as the background universe. The resulting p values were corrected for multiple testing using the Benjamini-Hochberg procedure and enrichments with $FDR <= 0.1$ are reported. These operations were performed in R using the HypeR package in R(*17*).

#### 3.4 Risk gene enrichment

For each disorder, a set of known risk genes were extracted from public databases as follows. For ASD, we obtained the Gene Scoring module from the SFARI Gene database(*18*). This module offers a comprehensive scoring indicating the strength of literature evidence supporting each gene's association with autism. We filtered the list to retain only genes labeled as 'Category 1' and 'Syndromic', as high confidence autism risk genes. For BPD and SCZ, we obtained disease gene associations listed in the DisGeneNet database(*19*). For both disorders, we retained only those genes that are labeled as "CTD_human" and used them for enrichment analysis.

#### 3.5 Enrichment of differentially expressed genes

To calculate the enrichment of differentially expressed genes within modules, we obtained the summary statistics for whole cortex differential gene expression results from ASD versus control from a previously published paper(*20*) (reported file Supplementary Data 3 in the publication). The log fold change values were used as a parameter. Briefly, the parametric analysis of gene set enrichment (PAGE) is a statistical method used to determine whether predefined sets of genes show statistically significant, coordinated differences in expression in different biological conditions or treatments(*21*). Unlike non-parametric methods like GSEA, which rank genes based on their expression levels and use permutation testing, PAGE employs a parametric approach that relies on the normal distribution to model gene expression changes. PAGE computes a Z-score for each gene set (module) based on the mean fold change of the genes within the set. The Z-score indicates how far the mean of the gene set is from the overall mean expression changes in units of standard deviation.

$$Z = \frac{\bar{d} - \mu_d}{\sigma_d}$$

where $\bar{d}$ is the mean expression change of the module, $\mu_d$ is the overall mean of all genes, and $\sigma_d$ is the standard deviation across all fold change values. The resulting Z-scores are shown in main text Figure 3.

### 4 Graph neural networks predict novel cell-type disorder genes

#### 4.1 Label creation for supervised learning

We created two sets of training labels for disorder cell type gene prioritization models for the purposes of comparison. For model 1, we used known disorder risk genes from public databases (**described in section 3.4**) as positive labels. Specifically, for ASD, we used candidate gene nominations within category gene.score = 1 or syndromic = 1 in the SFARI database. For SCZ and BPD, we selected genes within the DisGeNet database labeled as "CTD_human". Note that these sets are devoid of cell type specificity. For model 2, we appended model 1 labels with genes in modules enriched with known risk genes, thereby leveraging risk neighborhoods of the networks.

#### 4.2 Model training and evaluation

The model consists of a standard graph convolutional network (GCN) with 2 hidden layers and an output of dimension 2 for $n$ genes. Batch normalization layers are included after each hidden layer and immediately followed by Leaky ReLU activation functions. Typically, the dimension of the first and second hidden layers are set to 32 and 16, respectively. We refer to the first three layers as EMB and the last as CLASS. As we use no node features, the input, $X_i$ for each gene $i$ is equal to a zero-array of length $n$ with 1 at position $i$. Each model has an additional FCL to predict the weight of a directed edge given embeddings corresponding to each gene. The models each use two losses: (1) Embedding loss, which is the MSE of the predicted adjacency matrix by the model compared to the actual adjacency matrix and (2) classification loss, which is the cross entropy loss between the model output layer and the true labels. These may also be represented as

$$\mathrm{l}_{\mathrm{Embedding}} = \frac{1}{n^2}\sum_{i=1}^{n}\sum_{j=1, j\neq i}^{n} \mathrm{FCL}\left(\mathrm{EMB}(X_i), \mathrm{EMB}(X_j)\right) - X_{ij} \quad (1)$$

$$\sum_{i=1}^{n} Y_i \cdot \log \mathrm{CLASS}\left(\mathrm{GCN}(X_i)\right) + (1 - Y_i) \cdot \log \mathrm{CLASS}\left(\mathrm{GCN}(X_i)\right) \quad (2)$$

Where $n$ is the number of genes, $\mathrm{embedding}_i$ is the embedding associated with gene $i$, and $X_{ij}$ is the weight of the edge between genes $i$ and $j$. The two losses iterate the model separately, with the embedding layers frozen for the backpropagation of the classification loss.

Each cell type is trained individually using five fold cross-validation. More precisely, the data is split into 5 partitions (folds) and a model is trained using each partition as validation data, resulting in 5 separate models. The mean and standard deviation of the scores for each gene are then recorded and taken as the final model predictions. The folds are analyzed individually for performance statistics such as AUROC and AUPRC.

#### 4.3 BBB Drug prioritization

For each cell type, we examine the top decile of genes prioritized within the model. Then, the resulting gene list is scanned for gene targets of drugs which were earlier predicted to cross the blood brain barrier using ML models(*10*). These counts are then shown in **Figure 4c** for both models 1 and 2.

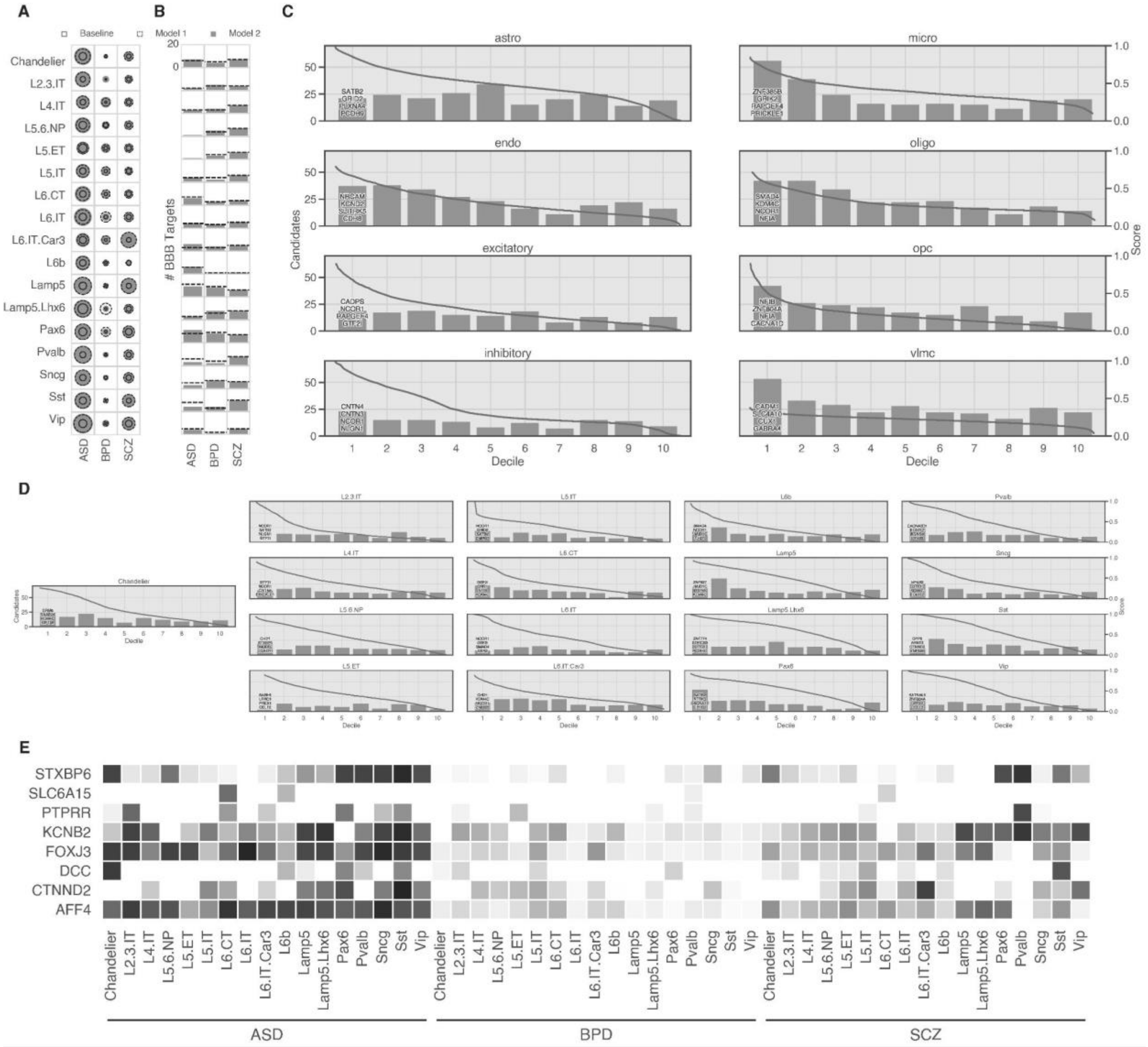


**Figure S7: Graph learning model prioritizes existing candidate disease genes and novel risk genes in ASD, BPD, and SCZ over precise cell type groupings. A)** Performance of the model with specified targets. Circle sizes scale from radii of 0-1 based on AUPRC for baseline (solid line), model 1 (disease genes only, dashed line), and model 2 (disease and module genes, filled circle). **B)** Number of targets of blood-brain barrier (BBB) drugs found in the top decile of prioritized genes by model 1 (dashed line) and model 2 (filled bar) from 0 to 20. **C)** Histogram of the number of SFARI candidate category 2 genes found in each decile of ASD predicting models for primary cell types. The top decile is annotated with the top 4 prioritized candidate genes. The model score (right axis) is also plotted against the prediction percentile (gray line). **D)** Repeated histogram plots for subclass cell types. **E)** Top 3 prioritized genes from each disease and their respective prioritizations within each cell type. Missing entries were not included in the corresponding cell type GRN. Cell types are categorized by disease and all scores are shaded based on per-model normalized risk scores. Darker squares indicate higher model score.

### 5 Network-based drug repurposing

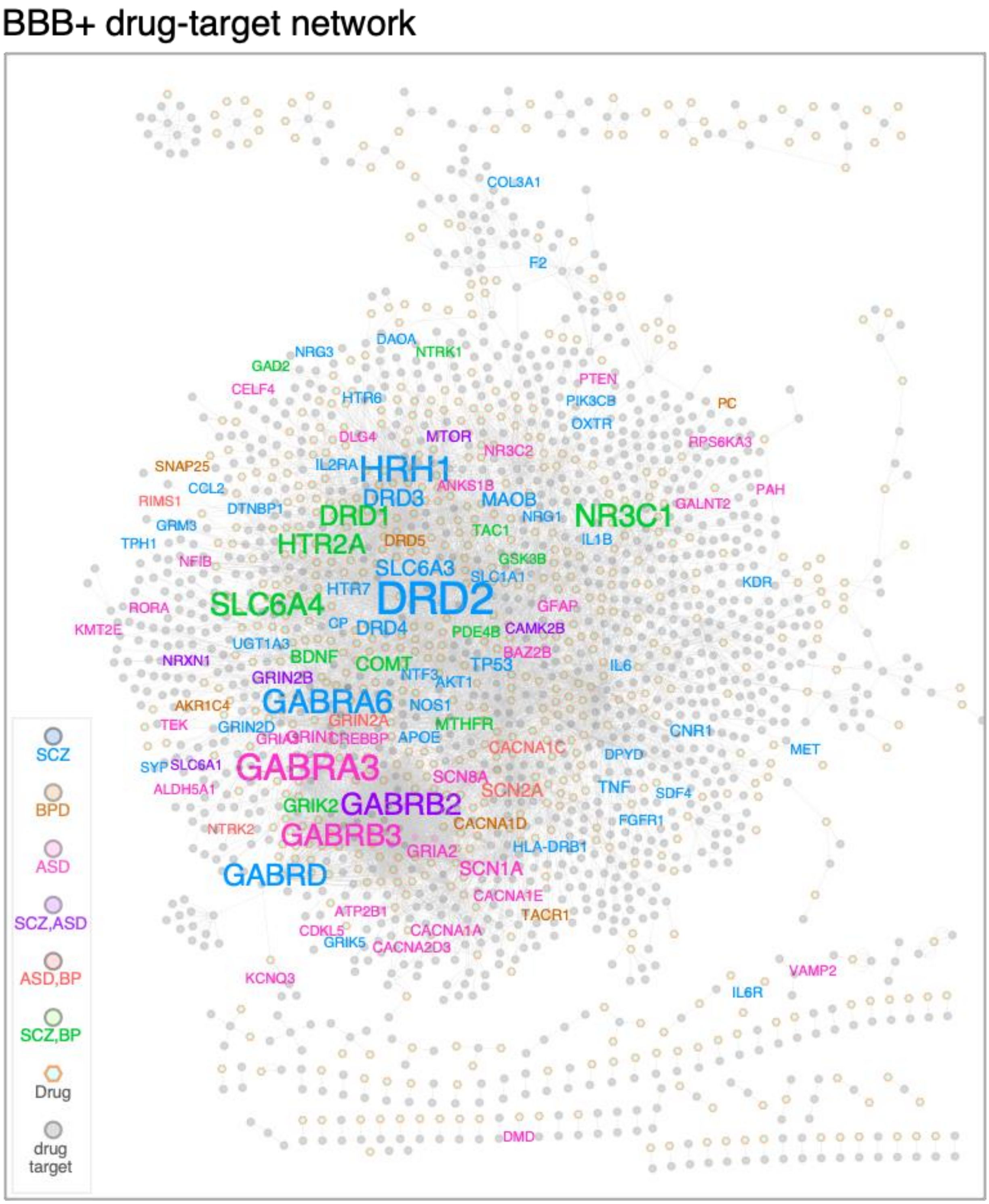


**Figure S8: BBB permeable drug target network.** The network was created by integrating drug-target links in the DrugBank database and the Drug-Gene Interaction Database. Drugs there are listed as BBB permeable (gray circles) are retained and rest filtered. Disorder risk genes (SFARI database for ASD and

DisGenNet for BPD and SCZ) are mapped onto the BBB+ drug targets and colored uniquely as shown in the legend. Genes that are targets of BBB+ drugs but not in the list of risk genes are shown in white circle. The node and label sizes are set proportional to the indegree, with genes targeted by several drugs shown larger relative to those that are targeted by a single drug. Labels for drug nodes and non-risk genes are kept hidden for ease in visualization.

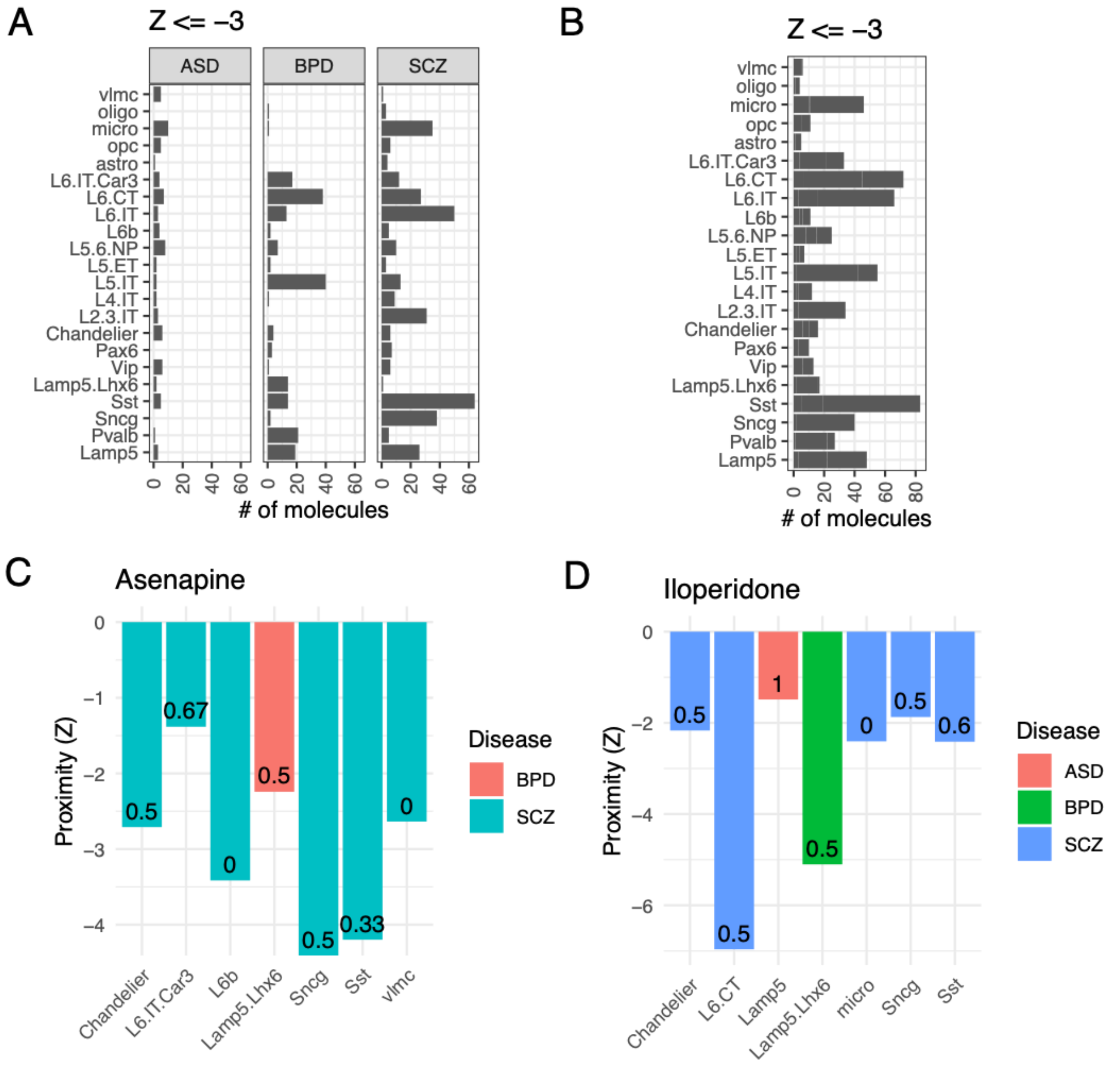


**Figure S9. Network based drug repurposing.** A) A plot showing number of molecules (x-axis) found significantly proximal to disorder risk genes within cell type networks (y-axis) across the three disorders (facets). B) The plot shows the number of molecules found significantly proximal to risk genes within cell type networks across all disorders (y-axis). C-D) Example drug molecules correctly recovered within our drug repurposing pipeline. The Z scores of network proximity is shown on the y-axis and cell types on the x-axis. The bars are colored uniquely for each disorder as shown in the legend.

#### 5.1 BBB permeable drug-target network

We acquired an academic license for the DrugBank database and extracted all drug IDs along with their corresponding target genes. Additionally, we obtained data from the Drug-Gene Interaction Database (DGIDB)(*22*), from which we filtered out compounds classified as "approved == TRUE" and "anti_neoplastic == FALSE." Concurrently, we obtained blood-brain-barrier (BBB) permeability labels for small molecules from the B3DB(*10*). The B3DB contains includes both numerical (log BB) and categorical (BBB+ or BBB−) data for over 7,800 compounds, sourced from over 50 publications. It contains both quantitative (log BB values) and qualitative (BBB+ or BBB−) measurements for drug compounds. The database is designed to aid machine learning models for predicting BBB permeability, addressing limitations of smaller, less diverse datasets and making it a valuable resource for drug discovery targeting central nervous system diseases. We matched the IUPAC names of BBB+ molecules with those listed in DrugBank to extract the drug IDs for brain disorders. We cross-referenced the names of BBB+ molecules with the *drug_claim_name* entries in DGIDB and combined them with the BBB+ compounds from DrugBank to create a network of 802 drugs along with their 1196 target genes for further analysis, as described below.

#### 5.2 Network proximity

We leverage the relationships between drugs, their targets, and disease-associated genes within a cell type network for drug repurposing. This approach to identify potential drug candidates that can be repurposed to treat diseases based on their "proximity" to disorder-associated nodes within a network graph. To create a pool of disease/disorder specific nodes, we utilized novel predictions from our GNN models (model 2 described in section 4) along with the positive training lables. This allows us to use cell type relevant risk genes to augment current knowledge on the genes underlying the disorder. Further, we used cell type coregulatory networks **(described in section 3.1**) as graphs for drug repurposing instead of using bipartite GRNs to avoid degree and connectivity biases.

To analyze the network proximity between drug targets and disorder genes in a coregulatory network, we compute various statistics to assess whether drug targets are closer to disorder genes than would be expected by chance. First, we calculate the degree distribution of both drug targets and disorder genes, which represents the number of connections each node has in the network. This is important for ensuring that our random selection of nodes for baseline comparison maintains similar connectivity properties. Next, we calculate the proximity between two sets of nodes (drug targets and disorder genes) based on their shortest path distances in the network. We compute the mean of the minimum distances between each pair of nodes from drug targets to disorder genes. To create a baseline for comparison, we compute a random distribution of proximity values by randomly selecting nodes from the network that match the degree distribution of the actual drug targets and disorder genes. We calculate their proximity iteratively 100 times and use this random distribution as a baseline to assess the statistical significance of the observed proximity. We compute a Z-score and a p-value of the observed proximity. All observations with a p-value < 0.1 are reported. All these operations were performed using the igraph library in R (https://igraph.org).

#### 5.3 Reversers vs mimickers

We obtained the Library of Integrated Network-Based Cellular Signatures (LINCS) dataset from SigCom(*8*). Specifically, we worked with the L1000 Chemical Perturbations dataset that provides up and down regulated gene sets in response to chemical perturbations measured across several human cell lines. We extracted experiments conducted in CNS cell lines (“NEU” cell lines) with the repurposable drug molecules identified in our study. In parallel, we obtained results from the differential gene expression (DGE) analysis for the disorders reported by us previously (*1*). Then, for the set of up- and- down-regulated genes for each repurposable drug in the L1000 data, we calculated the mean fold change in disorder. We then compared the direction of changes and labeled drugs as ‘reverser’ if the mean is in the opposite direction to that reported in the LINCS1000 dataset for both up- and- down- sets for the drug, or ‘mimiker’ otherwise.

### 6 Drug-associated cell-type eQTLs

We carried out a set of QTL analyses to identify cases for which genotypic variation is associated with significant changes in expression for the gene sets targeted by different drugs. This entailed using a total sample size of 125 (after filtering out duplicate samples or samples with missing covariates, and by including only those samples for which genotype PCs had been calculated in (*1*) As the response variable in this QTL analysis, for a given drug found to be repurposable in our analysis, we first identified the set of genes which are known to be affected by that drug. For a given drug, we refer to this set of genes as the drug’s “drug target genes”. Then, for each donor, we log-normalized the gene expression matrix and calculated the average expression of the drug’s drug target genes within each cell type. These average expression values, for every drug-cell type combination, were then used to populate a 'DrugCell' matrix (here, the term ‘DrugCell’ is adopted as a short term for a drug-cell type combination). In this matrix, rows represented drug-cell combinations, and columns corresponded to donors. The DrugCell matrix was then used as input to eQTL analysis.

We used QTLtools (version 1.3.1)(*23*) and ran our calculations in nominal-pass mode. Because each DrugCell comprises multiple genes, the nominal-pass calculation was performed by placing each DrugCell in each of the 22 autosomal human chromosomes, and then performing a search for SNPs within a cis-window that engulfs each entire chromosome in full. We used biological sex, age of death, diagnosis, cohort, five genotype PCs (to control for ancestry), and 5 expression PCs (to control for hidden batch effects) as covariates in our analysis. We filtered out low-frequency variants by removing any variant with a minor allele frequency (MAF) of less than 0.05. This yielded a total of 1,953,349 SNPs.

Specifically, we used the following format as the command when running QTLtools (again, the large cis window was used to ensure that our calculations covered the entirety of each chromosome):

*QTLtools cis --vcf vcf_file_containing_genotype_variants --bed bed_file_containing_drugcell_phenotype_values --cov matrix_containing_covariates --nominal 2.5597064323886822e-08 --window 300000000 --normal --out output_file_name*

Thus, in sum, this analysis is carried out by performing linear regression, wherein we included covariates as well as the response variable (i.e., DrugCell values) in a linear regression on genotype dosage. The residual DrugCell values were rank normal transformed (after the covariate correction). In order to control for the extremely large number of tests for each individual DrugCell (namely, the number of filtered SNPs), P-values in this analysis were corrected using Bonferroni correction at the level of 0.05, with this correction having been applied at the level of each DrugCell separately (thus, the nominal p-value threshold was ~2.56e-08 = 0.05/1,953,349). This yielded a total of 335 significant Drug-associated cell-type eQTLs, and we make these available as supplementary data file S10. The format for this output file is given as follows:

1. The drugCell name
2. The variant ID
3. The variant chromosome
4. The start position of the variant
5. The nominal p-value of the association between the variant and the DrugCell
6. The r2 of the linear regression
7. The beta (slope) of the linear regression

### 7 Development of psyDGN

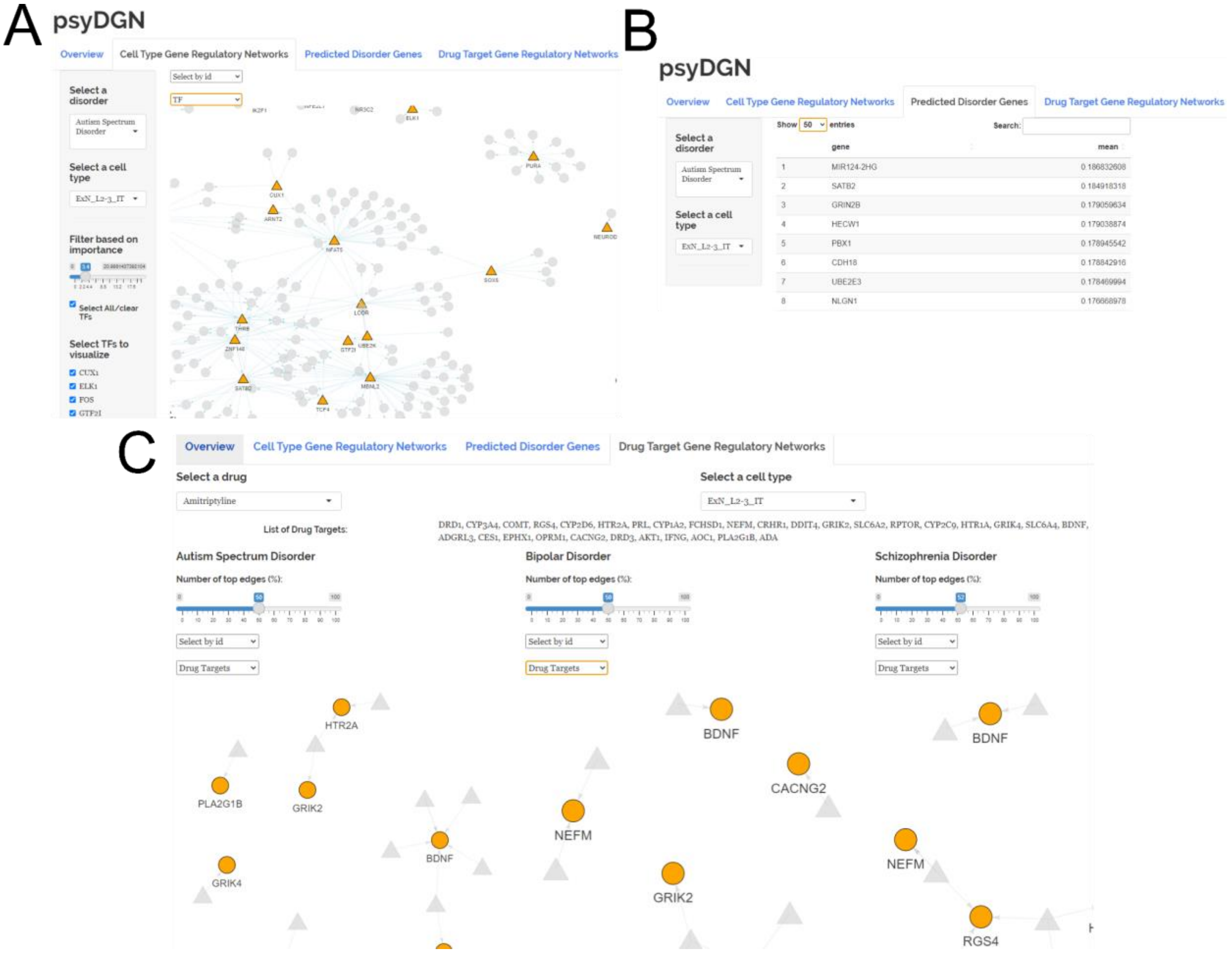


**Figure S10: Screen shots of the psyDGN web application.**

We developed an R shiny web application, psyDGN (link xx), allowing users to interactively explore the findings of the current study. The psyDGN consists of three sub-applications. In the first sub-application, it allows users to visualize cell-type gene regulatory networks (GRNs). Users can select one of the three disorders (Autism spectrum disorder, bipolar disorder, and Schizophrenia) and a cell type to visualize a corresponding disease-cell-type GRN. Users can also filter GRNs based on the edge importance scores and isolate connections specific to selected transcription factors. In the second sub-application, users can tabulate the predicted disorder genes for a given cell type and a disorder. In the third sub-application, users can visualize cell type GRN overlaps with drug targets. The psyDGN allows users to select a drug and a cell type of their choice from a drop-down menu and outputs three networks corresponding to the three disorders. Only the transcription factor-target gene connections overlapped with the drug-target genes are visualized.

**Supplementary Data:**

Supplementary Data S1: List of TFs that occur in the top 10% betweenness and outdegrees across cell types and disorders.

Supplementary Data S2: Disease influence scores of TFs across three disorders.

Supplementary Data S3: List of TFs with largest disease influence scores along with their expression patterns in response to BBB permeable drugs in LINCS dataset.
Supplementary Data S4: List of genes and their corresponding modules across cell types and disorders.
Supplementary Data S5: GO BP enrichment analysis of coregulated modules.
Supplementary Data S6: Risk gene prediction results.
Supplementary Data S7: The BBB permeable drug-target network curated from public databases and used for drug repurposing in our analysis.
Supplementary Data S8: Results of the drug proximity analysis.
Supplementary Data S9: List of mimickers and reversers.
Supplementary Data S10: Results of the drug-associated cell type eQTL analysis.

**References**


1. P. S. Emani, J. J. Liu, D. Clarke, M. Jensen, J. Warrell, C. Gupta, R. Meng, C. Y. Lee, S. Xu, C. Dursun, S. Lou, Y. Chen, Z. Chu, T. Galeev, A. Hwang, Y. Li, P. Ni, X. Zhou, PsychENCODE Consortium‡, T. E. Bakken, J. Bendl, L. Bicks, T. Chatterjee, L. Cheng, Y. Cheng, Y. Dai, Z. Duan, M. Flaherty, J. F. Fullard, M. Gancz, D. Garrido-Martín, S. Gaynor-Gillett, J. Grundman, N. Hawken, E. Henry, G. E. Hoffman, A. Huang, Y. Jiang, T. Jin, N. L. Jorstad, R. Kawaguchi, S. Khullar, J. Liu, J. Liu, S. Liu, S. Ma, M. Margolis, S. Mazariegos, J. Moore, J. R. Moran, E. Nguyen, N. Phalke, M. Pjanic, H. Pratt, D. Quintero, A. S. Rajagopalan, T. R. Riesenmy, N. Shedd, M. Shi, M. Spector, R. Terwilliger, K. J. Travaglini, B. Wamsley, G. Wang, Y. Xia, S. Xiao, A. C. Yang, S. Zheng, M. J. Gandal, D. Lee, E. S. Lein, P. Roussos, N. Sestan, Z. Weng, K. P. White, H. Won, M. J. Girgenti, J. Zhang, D. Wang, D. Geschwind, M. Gerstein, PsychENCODE Consortium, Single-cell genomics and regulatory networks for 388 human brains. *Science* **384**, eadi5199 (2024).

2. Y. Hao, T. Stuart, M. H. Kowalski, S. Choudhary, P. Hoffman, A. Hartman, A. Srivastava, G. Molla, S. Madad, C. Fernandez-Granda, R. Satija, Dictionary learning for integrative, multimodal and scalable single-cell analysis. *Nat Biotechnol* **42**, 293–304 (2024).

3. F. A. Wolf, P. Angerer, F. J. Theis, SCANPY: large-scale single-cell gene expression data analysis. *Genome Biol* **19**, 15 (2018).

4. R. Janky, A. Verfaillie, H. Imrichová, B. Van de Sande, L. Standaert, V. Christiaens, G. Hulselmans, K. Herten, M. Naval Sanchez, D. Potier, D. Svetlichnyy, Z. Kalender Atak, M. Fiers, J.-C. Marine, S. Aerts, iRegulon: from a gene list to a gene regulatory network using large motif and track collections. *PLoS Comput Biol* **10**, e1003731 (2014).

5. T. Moerman, S. Aibar Santos, C. Bravo González-Blas, J. Simm, Y. Moreau, J. Aerts, S. Aerts, GRNBoost2 and Arboreto: efficient and scalable inference of gene regulatory networks. *Bioinformatics* **35**, 2159–2161 (2019).

6. S. Aibar, C. B. González-Blas, T. Moerman, V. A. Huynh-Thu, H. Imrichova, G. Hulselmans, F. Rambow, J.-C. Marine, P. Geurts, J. Aerts, J. van den Oord, Z. K. Atak, J. Wouters, S. Aerts, SCENIC: single-cell regulatory network inference and clustering. *Nat Methods* **14**, 1083–1086 (2017).

7. P. S. Emani, J. J. Liu, D. Clarke, M. Jensen, J. Warrell, C. Gupta, R. Meng, C. Y. Lee, S. Xu, C. Dursun, S. Lou, Y. Chen, Z. Chu, T. Galeev, A. Hwang, Y. Li, P. Ni, X. Zhou, PsychENCODE Consortium, T. E. Bakken, J. Bendl, L. Bicks, T. Chatterjee, L. Cheng, Y. Cheng, Y. Dai, Z. Duan, M. Flaherty, J. F. Fullard, M. Gancz, D. Garrido-Martín, S. Gaynor-Gillett, J. Grundman, N. Hawken, E. Henry, G. E. Hoffman, A. Huang, Y. Jiang, T. Jin, N. L. Jorstad, R. Kawaguchi, S. Khullar, J. Liu, J. Liu, S. Liu, S. Ma, M. Margolis, S. Mazariegos, J. Moore, J. R. Moran, E. Nguyen, N. Phalke, M. Pjanic, H. Pratt, D. Quintero, A. S. Rajagopalan, T. R. Riesenmy, N. Shedd, M. Shi, M. Spector, R. Terwilliger, K. J. Travaglini, B. Wamsley, G. Wang, Y. Xia, S. Xiao, A. C. Yang, S. Zheng, M. J.

Gandal, D. Lee, E. S. Lein, P. Roussos, N. Sestan, Z. Weng, K. P. White, H. Won, M. J. Girgenti, J. Zhang, D. Wang, D. Geschwind, M. Gerstein, Single-cell genomics and regulatory networks for 388 human brains. *bioRxiv*, 2024.03.18.585576 (2024).

8. J. E. Evangelista, D. J. B. Clarke, Z. Xie, A. Lachmann, M. Jeon, K. Chen, K. M. Jagodnik, S. L. Jenkins, M. V. Kuleshov, M. L. Wojciechowicz, S. C. Schürer, M. Medvedovic, A. Ma'ayan, SigCom LINCS: data and metadata search engine for a million gene expression signatures. *Nucleic Acids Res* **50**, W697–W709 (2022).

9. Q. Duan, S. P. Reid, N. R. Clark, Z. Wang, N. F. Fernandez, A. D. Rouillard, B. Readhead, S. R. Tritsch, R. Hodos, M. Hafner, M. Niepel, P. K. Sorger, J. T. Dudley, S. Bavari, R. G. Panchal, A. Ma'ayan, L1000CDS2: LINCS L1000 characteristic direction signatures search engine. *NPJ Syst Biol Appl* **2**, 16015- (2016).

10. F. Meng, Y. Xi, J. Huang, P. W. Ayers, A curated diverse molecular database of blood-brain barrier permeability with chemical descriptors. *Sci Data* **8**, 289 (2021).

11. P. Langfelder, S. Horvath, WGCNA: an R package for weighted correlation network analysis. *BMC Bioinformatics* **9**, 559 (2008).

12. V. Trubetskoy, A. F. Pardiñas, T. Qi, G. Panagiotaropoulou, S. Awasthi, T. B. Bigdeli, J. Bryois, C.-Y. Chen, C. A. Dennison, L. S. Hall, M. Lam, K. Watanabe, O. Frei, T. Ge, J. C. Harwood, F. Koopmans, S. Magnusson, A. L. Richards, J. Sidorenko, Y. Wu, J. Zeng, J. Grove, M. Kim, Z. Li, G. Voloudakis, W. Zhang, M. Adams, I. Agartz, E. G. Atkinson, E. Agerbo, M. Al Eissa, M. Albus, M. Alexander, B. Z. Alizadeh, K. Alptekin, T. D. Als, F. Amin, V. Arolt, M. Arrojo, L. Athanasiu, M. H. Azevedo, S. A. Bacanu, N. J. Bass, M. Begemann, R. A. Belliveau, J. Bene, B. Benyamin, S. E. Bergen, G. Blasi, J. Bobes, S. Bonassi, A. Braun, R. A. Bressan, E. J. Bromet, R. Bruggeman, P. F. Buckley, R. L. Buckner, J. Bybjerg-Grauholm, W. Cahn, M. J. Cairns, M. E. Calkins, V. J. Carr, D. Castle, S. V. Catts, K. D. Chambert, R. C. K. Chan, B. Chaumette, W. Cheng, E. F. C. Cheung, S. A. Chong, D. Cohen, A. Consoli, Q. Cordeiro, J. Costas, C. Curtis, M. Davidson, K. L. Davis, L. de Haan, F. Degenhardt, L. E. DeLisi, D. Demontis, F. Dickerson, D. Dikeos, T. Dinan, S. Djurovic, J. Duan, G. Ducci, F. Dudbridge, J. G. Eriksson, L. Fañanás, S. V. Faraone, A. Fiorentino, A. Forstner, J. Frank, N. B. Freimer, M. Fromer, A. Frustaci, A. Gadelha, G. Genovese, E. S. Gershon, M. Giannitelli, I. Giegling, P. Giusti-Rodríguez, S. Godard, J. I. Goldstein, J. González Peñas, A. González-Pinto, S. Gopal, J. Gratten, M. F. Green, T. A. Greenwood, O. Guillin, S. Gülöksüz, R. E. Gur, R. C. Gur, B. Gutiérrez, E. Hahn, H. Hakonarson, V. Haroutunian, A. M. Hartmann, C. Harvey, C. Hayward, F. A. Henskens, S. Herms, P. Hoffmann, D. P. Howrigan, M. Ikeda, C. Iyegbe, I. Joa, A. Julià, A. K. Kähler, T. Kam-Thong, Y. Kamatani, S. Karachanak-Yankova, O. Kebir, M. C. Keller, B. J. Kelly, A. Khrunin, S.-W. Kim, J. Klovins, N. Kondratiev, B. Konte, J. Kraft, M. Kubo, V. Kučinskas, Z. A. Kučinskiene, A. Kusumawardhani, H. Kuzelova-Ptackova, S. Landi, L. C. Lazzeroni, P. H. Lee, S. E. Legge, D. S. Lehrer, R. Lencer, B. Lerer, M. Li, J. Lieberman, G. A. Light, S. Limborska, C.-M. Liu, J. Lönnqvist, C. M. Loughland, J. Lubinski, J. J. Luykx, A. Lynham, M. Macek, A. Mackinnon, P. K. E. Magnusson, B. S. Maher, W. Maier, D. Malaspina, J. Mallet, S. R. Marder, S. Marsal, A. R. Martin, L. Martorell, M. Mattheisen, R. W. McCarley, C. McDonald, J. J. McGrath, H. Medeiros, S. Meier, B. Melegh, I. Melle, R. I. Mesholam-Gately, A. Metspalu, P. T. Michie, L. Milani, V. Milanova, M. Mitjans, E. Molden, E. Molina, M. D. Molto, V. Mondelli, C. Moreno, C. P. Morley, G. Muntané, K. C. Murphy, I. Myin-Germeys, I. Nenadić, G. Nestadt, L. Nikitina-Zake, C. Noto, K. H. Nuechterlein, N. L. O'Brien, F. A. O'Neill, S.-Y. Oh, A. Olincy, V. K. Ota, C. Pantelis, G. N. Papadimitriou, M. Parellada, T. Paunio, R. Pellegrino, S. Periyasamy, D. O. Perkins, B. Pfuhlmann, O. Pietiläinen, J. Pimm, D. Porteous, J. Powell, D. Quattrone, D. Quested, A. D. Radant, A. Rampino, M. H. Rapaport, A. Rautanen, A. Reichenberg, C. Roe, J. L. Roffman, J. Roth, M. Rothermundt, B. P. F. Rutten, S. Saker-Delye, V. Salomaa, J. Sanjuan, M. L. Santoro, A. Savitz, U. Schall, R. J. Scott, L. J. Seidman, S. I. Sharp, J. Shi, L. J. Siever, E. Sigurdsson, K. Sim, N. Skarabis, P. Slominsky, H.-C. So, J. L. Sobell, E. Söderman, H. J. Stain, N. E. Steen, A. A. Steixner-Kumar, E. Stögmann, W. S. Stone, R. E. Straub, F. Streit, E. Strengman, T. S. Stroup, M. Subramaniam, C. A. Sugar, J. Suvisaari, D. M. Svrakic, N. R. Swerdlow, J. P. Szatkiewicz, T. M. T.

Ta, A. Takahashi, C. Terao, F. Thibaut, D. Toncheva, P. A. Tooney, S. Torretta, S. Tosato, G. B. Tura, B. I. Turetsky, A. Üçok, A. Vaaler, T. van Amelsvoort, R. van Winkel, J. Veijola, J. Waddington, H. Walter, A. Waterreus, B. T. Webb, M. Weiser, N. M. Williams, S. H. Witt, B. K. Wormley, J. Q. Wu, Z. Xu, R. Yolken, C. C. Zai, W. Zhou, F. Zhu, F. Zimprich, E. C. Atbaşoğlu, M. Ayub, C. Benner, A. Bertolino, D. W. Black, N. J. Bray, G. Breen, N. G. Buccola, W. F. Byerley, W. J. Chen, C. R. Cloninger, B. Crespo-Facorro, G. Donohoe, R. Freedman, C. Galletly, M. J. Gandal, M. Gennarelli, D. M. Hougaard, H.-G. Hwu, A. V. Jablensky, S. A. McCarroll, J. L. Moran, O. Mors, P. B. Mortensen, B. Müller-Myhsok, A. L. Neil, M. Nordentoft, M. T. Pato, T. L. Petryshen, M. Pirinen, A. E. Pulver, T. G. Schulze, J. M. Silverman, J. W. Smoller, E. A. Stahl, D. W. Tsuang, E. Vilella, S.-H. Wang, S. Xu, Indonesia Schizophrenia Consortium, PsychENCODE, Psychosis Endophenotypes International Consortium, SynGO Consortium, R. Adolfsson, C. Arango, B. T. Baune, S. I. Belangero, A. D. Børglum, D. Braff, E. Bramon, J. D. Buxbaum, D. Campion, J. A. Cervilla, S. Cichon, D. A. Collier, A. Corvin, D. Curtis, M. D. Forti, E. Domenici, H. Ehrenreich, V. Escott-Price, T. Esko, A. H. Fanous, A. Gareeva, M. Gawlik, P. V. Gejman, M. Gill, S. J. Glatt, V. Golimbet, K. S. Hong, C. M. Hultman, S. E. Hyman, N. Iwata, E. G. Jönsson, R. S. Kahn, J. L. Kennedy, E. Khusnutdinova, G. Kirov, J. A. Knowles, M.-O. Krebs, C. Laurent-Levinson, J. Lee, T. Lencz, D. F. Levinson, Q. S. Li, J. Liu, A. K. Malhotra, D. Malhotra, A. McIntosh, A. McQuillin, P. R. Menezes, V. A. Morgan, D. W. Morris, B. J. Mowry, R. M. Murray, V. Nimgaonkar, M. M. Nöthen, R. A. Ophoff, S. A. Paciga, A. Palotie, C. N. Pato, S. Qin, M. Rietschel, B. P. Riley, M. Rivera, D. Rujescu, M. C. Saka, A. R. Sanders, S. G. Schwab, A. Serretti, P. C. Sham, Y. Shi, D. St Clair, H. Stefánsson, K. Stefansson, M. T. Tsuang, J. van Os, M. P. Vawter, D. R. Weinberger, T. Werge, D. B. Wildenauer, X. Yu, W. Yue, P. A. Holmans, A. J. Pocklington, P. Roussos, E. Vassos, M. Verhage, P. M. Visscher, J. Yang, D. Posthuma, O. A. Andreassen, K. S. Kendler, M. J. Owen, N. R. Wray, M. J. Daly, H. Huang, B. M. Neale, P. F. Sullivan, S. Ripke, J. T. R. Walters, M. C. O'Donovan, Schizophrenia Working Group of the Psychiatric Genomics Consortium, Mapping genomic loci implicates genes and synaptic biology in schizophrenia. *Nature* **604**, 502–508 (2022).

13. J. Grove, S. Ripke, T. D. Als, M. Mattheisen, R. K. Walters, H. Won, J. Pallesen, E. Agerbo, O. A. Andreassen, R. Anney, S. Awashti, R. Belliveau, F. Bettella, J. D. Buxbaum, J. Bybjerg-Grauholm, M. Bækvad-Hansen, F. Cerrato, K. Chambert, J. H. Christensen, C. Churchhouse, K. Dellenvall, D. Demontis, S. De Rubeis, B. Devlin, S. Djurovic, A. L. Dumont, J. I. Goldstein, C. S. Hansen, M. E. Hauberg, M. V. Hollegaard, S. Hope, D. P. Howrigan, H. Huang, C. M. Hultman, L. Klei, J. Maller, J. Martin, A. R. Martin, J. L. Moran, M. Nyegaard, T. Nærland, D. S. Palmer, A. Palotie, C. B. Pedersen, M. G. Pedersen, T. dPoterba, J. B. Poulsen, B. S. Pourcain, P. Qvist, K. Rehnström, A. Reichenberg, J. Reichert, E. B. Robinson, K. Roeder, P. Roussos, E. Saemundsen, S. Sandin, F. K. Satterstrom, G. Davey Smith, H. Stefansson, S. Steinberg, C. R. Stevens, P. F. Sullivan, P. Turley, G. B. Walters, X. Xu, Autism Spectrum Disorder Working Group of the Psychiatric Genomics Consortium, BUPGEN, Major Depressive Disorder Working Group of the Psychiatric Genomics Consortium, 23andMe Research Team, K. Stefansson, D. H. Geschwind, M. Nordentoft, D. M. Hougaard, T. Werge, O. Mors, P. B. Mortensen, B. M. Neale, M. J. Daly, A. D. Børglum, Identification of common genetic risk variants for autism spectrum disorder. *Nat Genet* **51**, 431–444 (2019).

14. N. Mullins, A. J. Forstner, K. S. O'Connell, B. Coombes, J. R. I. Coleman, Z. Qiao, T. D. Als, T. B. Bigdeli, S. Børte, J. Bryois, A. W. Charney, O. K. Drange, M. J. Gandal, S. P. Hagenaars, M. Ikeda, N. Kamitaki, M. Kim, K. Krebs, G. Panagiotaropoulou, B. M. Schilder, L. G. Sloofman, S. Steinberg, V. Trubetskoy, B. S. Winsvold, H.-H. Won, L. Abramova, K. Adorjan, E. Agerbo, M. Al Eissa, D. Albani, N. Alliey-Rodriguez, A. Anjorin, V. Antilla, A. Antoniou, S. Awasthi, J. H. Baek, M. Bækvad-Hansen, N. Bass, M. Bauer, E. C. Beins, S. E. Bergen, A. Birner, C. Bøcker Pedersen, E. Bøen, M. P. Boks, R. Bosch, M. Brum, B. M. Brumpton, N. Brunkhorst-Kanaan, M. Budde, J. Bybjerg-Grauholm, W. Byerley, M. Cairns, M. Casas, P. Cervantes, T.-K. Clarke, C. Cruceanu, A. Cuellar-Barboza, J. Cunningham, D. Curtis, P. M. Czerski, A. M. Dale, N. Dalkner, F. S. David, F. Degenhardt, S. Djurovic, A. L. Dobbyn, A. Douzenis, T. Elvsåshagen, V. Escott-Price, I. N. Ferrier, A. Fiorentino, T. M. Foroud, L. Forty, J. Frank, O. Frei, N. B. Freimer, L. Frisén, K. Gade, J. Garnham, J. Gelernter, M. Giørtz Pedersen, I. R. Gizer, S. D. Gordon, K. Gordon-Smith, T. A. Greenwood, J. Grove, J. Guzman-Parra, K. Ha, M. Haraldsson, M. Hautzinger, U. Heilbronner, D.

Hellgren, S. Herms, P. Hoffmann, P. A. Holmans, L. Huckins, S. Jamain, J. S. Johnson, J. L. Kalman, Y. Kamatani, J. L. Kennedy, S. Kittel-Schneider, J. A. Knowles, M. Kogevinas, M. Koromina, T. M. Kranz, H. R. Kranzler, M. Kubo, R. Kupka, S. A. Kushner, C. Lavebratt, J. Lawrence, M. Leber, H.-J. Lee, P. H. Lee, S. E. Levy, C. Lewis, C. Liao, S. Lucae, M. Lundberg, D. J. MacIntyre, S. H. Magnusson, W. Maier, A. Maihofer, D. Malaspina, E. Maratou, L. Martinsson, M. Mattheisen, S. A. McCarroll, N. W. McGregor, P. McGuffin, J. D. McKay, H. Medeiros, S. E. Medland, V. Millischer, G. W. Montgomery, J. L. Moran, D. W. Morris, T. W. Mühleisen, N. O'Brien, C. O'Donovan, L. M. Olde Loohuis, L. Oruc, S. Papiol, A. F. Pardiñas, A. Perry, A. Pfennig, E. Porichi, J. B. Potash, D. Quested, T. Raj, M. H. Rapaport, J. R. DePaulo, E. J. Regeer, J. P. Rice, F. Rivas, M. Rivera, J. Roth, P. Roussos, D. M. Ruderfer, C. Sánchez-Mora, E. C. Schulte, F. Senner, S. Sharp, P. D. Shilling, E. Sigurdsson, L. Sirignano, C. Slaney, O. B. Smeland, D. J. Smith, J. L. Sobell, C. Søholm Hansen, M. Soler Artigas, A. T. Spijker, D. J. Stein, J. S. Strauss, B. Świątkowska, C. Terao, T. E. Thorgeirsson, C. Toma, P. Tooney, E.-E. Tsermpini, M. P. Vawter, H. Vedder, J. T. R. Walters, S. H. Witt, S. Xi, W. Xu, J. M. K. Yang, A. H. Young, H. Young, P. P. Zandi, H. Zhou, L. Zillich, HUNT All-In Psychiatry, R. Adolfsson, I. Agartz, M. Alda, L. Alfredsson, G. Babadjanova, L. Backlund, B. T. Baune, F. Bellivier, S. Bengesser, W. H. Berrettini, D. H. R. Blackwood, M. Boehnke, A. D. Børglum, G. Breen, V. J. Carr, S. Catts, A. Corvin, N. Craddock, U. Dannlowski, D. Dikeos, T. Esko, B. Etain, P. Ferentinos, M. Frye, J. M. Fullerton, M. Gawlik, E. S. Gershon, F. S. Goes, M. J. Green, M. Grigoroiu-Serbanescu, J. Hauser, F. Henskens, J. Hillert, K. S. Hong, D. M. Hougaard, C. M. Hultman, K. Hveem, N. Iwata, A. V. Jablensky, I. Jones, L. A. Jones, R. S. Kahn, J. R. Kelsoe, G. Kirov, M. Landén, M. Leboyer, C. M. Lewis, Q. S. Li, J. Lissowska, C. Lochner, C. Loughland, N. G. Martin, C. A. Mathews, F. Mayoral, S. L. McElroy, A. M. McIntosh, F. J. McMahon, I. Melle, P. Michie, L. Milani, P. B. Mitchell, G. Morken, O. Mors, P. B. Mortensen, B. Mowry, B. Müller-Myhsok, R. M. Myers, B. M. Neale, C. M. Nievergelt, M. Nordentoft, M. M. Nöthen, M. C. O'Donovan, K. J. Oedegaard, T. Olsson, M. J. Owen, S. A. Paciga, C. Pantelis, C. Pato, M. T. Pato, G. P. Patrinos, R. H. Perlis, D. Posthuma, J. A. Ramos-Quiroga, A. Reif, E. Z. Reininghaus, M. Ribasés, M. Rietschel, S. Ripke, G. A. Rouleau, T. Saito, U. Schall, M. Schalling, P. R. Schofield, T. G. Schulze, L. J. Scott, R. J. Scott, A. Serretti, C. Shannon Weickert, J. W. Smoller, H. Stefansson, K. Stefansson, E. Stordal, F. Streit, P. F. Sullivan, G. Turecki, A. E. Vaaler, E. Vieta, J. B. Vincent, I. D. Waldman, T. W. Weickert, T. Werge, N. R. Wray, J.-A. Zwart, J. M. Biernacka, J. I. Nurnberger, S. Cichon, H. J. Edenberg, E. A. Stahl, A. McQuillin, A. Di Florio, R. A. Ophoff, O. A. Andreassen, Genome-wide association study of more than 40,000 bipolar disorder cases provides new insights into the underlying biology. *Nat Genet* **53**, 817–829 (2021).

15. International League Against Epilepsy Consortium on Complex Epilepsies, Genome-wide mega-analysis identifies 16 loci and highlights diverse biological mechanisms in the common epilepsies. *Nat Commun* **9**, 5269 (2018).

16. M. Ashburner, C. A. Ball, J. A. Blake, D. Botstein, H. Butler, J. M. Cherry, A. P. Davis, K. Dolinski, S. S. Dwight, J. T. Eppig, M. A. Harris, D. P. Hill, L. Issel-Tarver, A. Kasarskis, S. Lewis, J. C. Matese, J. E. Richardson, M. Ringwald, G. M. Rubin, G. Sherlock, Gene ontology: tool for the unification of biology. The Gene Ontology Consortium. *Nat Genet* **25**, 25–29 (2000).

17. A. Federico, S. Monti, hypeR: an R package for geneset enrichment workflows. *Bioinformatics* **36**, 1307–1308 (2020).

18. B. S. Abrahams, D. E. Arking, D. B. Campbell, H. C. Mefford, E. M. Morrow, L. A. Weiss, I. Menashe, T. Wadkins, S. Banerjee-Basu, A. Packer, SFARI Gene 2.0: a community-driven knowledgebase for the autism spectrum disorders (ASDs). *Mol Autism* **4**, 36 (2013).

19. J. Piñero, À. Bravo, N. Queralt-Rosinach, A. Gutiérrez-Sacristán, J. Deu-Pons, E. Centeno, J. García-García, F. Sanz, L. I. Furlong, DisGeNET: a comprehensive platform integrating information on human disease-associated genes and variants. *Nucleic Acids Res* **45**, D833–D839 (2017).

20. M. J. Gandal, J. R. Haney, B. Wamsley, C. X. Yap, S. Parhami, P. S. Emani, N. Chang, G. T. Chen, G. D. Hoftman, D. de Alba, G. Ramaswami, C. L. Hartl, A. Bhattacharya, C. Luo, T. Jin, D. Wang, R.

Kawaguchi, D. Quintero, J. Ou, Y. E. Wu, N. N. Parikshak, V. Swarup, T. G. Belgard, M. Gerstein, B. Pasaniuc, D. H. Geschwind, Broad transcriptomic dysregulation occurs across the cerebral cortex in ASD. *Nature* **611**, 532–539 (2022).

21. S.-Y. Kim, D. J. Volsky, PAGE: parametric analysis of gene set enrichment. *BMC Bioinformatics* **6**, 144 (2005).

22. S. L. Freshour, S. Kiwala, K. C. Cotto, A. C. Coffman, J. F. McMichael, J. J. Song, M. Griffith, O. L. Griffith, A. H. Wagner, Integration of the Drug-Gene Interaction Database (DGIdb 4.0) with open crowdsource efforts. *Nucleic Acids Res* **49**, D1144–D1151 (2021).

23. O. Delaneau, H. Ongen, A. A. Brown, A. Fort, N. I. Panousis, E. T. Dermitzakis, A complete tool set for molecular QTL discovery and analysis. *Nat Commun* **8**, 15452 (2017).
